# Modifiable traits, healthy behaviours, and leucocyte telomere length

**DOI:** 10.1101/2021.12.13.21267707

**Authors:** Vasiliki Bountziouka, Crispin Musicha, Elias Allara, Stephen Kaptoge, Qingning Wang, Emanuele Di Angelantonio, Adam S Butterworth, John R Thompson, John N Danesh, Angela M Wood, Christopher P Nelson, Veryan Codd, Nilesh J Samani

**Affiliations:** Department of Cardiovascular Sciences, University of Leicester, Leicester, UK; NIHR Leicester Biomedical Research Centre, Glenfield Hospital, Leicester, UK; British Heart Foundation Cardiovascular Epidemiology Unit, Department of Public Health and Primary Care, University of Cambridge, Cambridge, UK; National Institute for Health Research Blood and Transplant Research Unit in Donor Health and Genomics, University of Cambridge, Cambridge, UK; British Heart Foundation Centre of Research Excellence, University of Cambridge, Cambridge, UK; Health Data Research UK Cambridge, Wellcome Genome Campus and University of Cambridge, Cambridge, UK; Department of Health Sciences, University of Leicester, Leicester, UK; Wellcome Sanger Institute, Wellcome Genome Campus, Hinxton, Cambridge, UK

## Abstract

**Background:** Telomere length is associated with risk of several age–related diseases and cancers. The extent to which telomere length may be modifiable through lifestyle and behaviour and whether this has any clinical consequences is unknown.

**Methods:** In up to 422,797 participants in UK Biobank, we investigated associations of leucocyte telomere length (LTL) with 117 potentially modifiable traits, as well as two indices of healthy behaviours incorporating smoking, physical activity, diet, maintenance of a healthy body weight and alcohol intake. Associations were interpreted as age–related change in LTL by dividing the trait beta coefficients with the age–coefficient. We used Mendelian Randomisation (MR) to test causality of the observed associations of educational attainment and smoking behaviour with LTL. We investigated whether the associations of LTL with 22 diseases were modified by the number of healthy behaviours and the extent to which the associations of more healthy behaviours with greater life expectancy and lower risk of coronary artery disease (CAD) may be mediated through LTL.

**Results:** 71 traits showed significant associations with LTL but most were modest, equivalent to <1 year of age–related change in LTL. In multivariable analyses of 17 traits with stronger associations (equivalent to ≥2 years of age–related change in LTL), five traits – oily fish intake, educational attainment, general health status, walking pace and current smoking – remained significant. MR analysis suggested that educational attainment and smoking behaviour causally affect LTL. Both indices of healthy behaviour were positively and linearly associated with LTL, with those with the healthiest behaviour having longer LTL equivalent to ∼3·5 years of age–related change in LTL when compared with those with the least heathy behaviours (P<0·001). However, healthy behaviours only explained <0·2% of the total variation in LTL and did not significantly modify the association of LTL with risk of any of the diseases studied. Neither the association of more healthy behaviours on greater life expectancy or lower risk of CAD were substantially mediated through LTL.

**Conclusions:** Several potentially modifiable traits and healthy behaviours have a quantifiable association with LTL, at least some of which are likely to be causal. However, these effects are not of a sufficient magnitude to substantially alter the association between LTL and various diseases or life expectancy.

## Introduction

Telomeres, the cap structures at the ends of chromosomes, vary in their average length between individuals, with telomere length (TL) being an important determinant of cellular replicative capacity.^1^ Inter–individual variation in TL, most commonly measured in blood (leucocyte telomere length, LTL) is associated with a range of human biological traits, diseases and lifespan, with growing evidence that some of these relationships are causal.^2–4^ LTL is a highly heritable trait with inter– individual variation both at birth and throughout the life course.^5–7^ Other major correlates of LTL are age, sex and ethnicity, with older age, male sex and white ethnicity associated with shorter LTL.^7^ The decrease in LTL with age in population studies has led to LTL being proposed as a marker of biological ageing.^8^ However, its relationship with age–related diseases is complex, with opposing relationships between shorter LTL and increased risk of degenerative diseases, such as coronary artery disease (CAD) and decreased risk of several cancers.^2–4^

The relationship between LTL and disease risk has led to much interest in understanding whether an individual can potentially modify their LTL and consequently the associated risk of disease. At the cellular level, TL shortens in response to oxidative and inflammatory stress.^9,10^ Many studies have therefore sought to explore the relationship between LTL and a range of potentially modifiable environmental and lifestyle factors associated with oxidative stress and inflammation, including smoking,^11,12^ obesity,^11,13^ diet^14,15^ and physical activity.^16,17^ However, many of these studies have shown conflicting results, perhaps in part due to moderate sample sizes. Meta–analyses have strengthened the evidence of associations between smoking and higher body mass index (BMI) and shorter LTL.^12,13^ Lower physical activity generally associates with shorter LTL, although studies remain inconsistent, potentially due to diversity of methods used in the assessment of physical activity.^16^ The relationship between LTL and dietary factors remains to be fully established although evidence suggests that a Mediterranean diet associates with longer LTL.^15^ Overall, the available studies suggest that healthy behaviours are potentially associated with longer LTL while unhealthy behaviours are associated with shorter LTL. In addition, other traits and behaviours that impact LTL await discovery.

UK Biobank (UKB, http://www.ukbiobank.ac.uk), is a large prospective study that recruited participants between 2006–2010 and has collected detailed and extensive information on lifestyle and behaviour.^18^ We have recently completed measurement of LTL in 474,074 participants in UK Biobank.^7^ We therefore sought to utilise this new resource to undertake a comprehensive analysis to identify potentially modifiable factors that might influence LTL, and also address the composite question of whether, and to what extent, healthy behaviour, defined using previously described indices,^19,20^ is associated with longer LTL. In addition, we investigated whether healthy behaviour influences the association of LTL with diseases and to what extent the associations of more healthy behaviours with greater life expectancy and lower risk of CAD may be mediated through LTL.

## Methods

### Participants and LTL measurements

LTL was measured on DNA extracted from blood samples collected at baseline using an established qPCR method and reported as a ratio of the telomere repeat number (T) to a single–copy gene (S) (T/S ratio). The measurements were loge–transformed to approximate the normal distribution and were then transformed to z–standardised values (UKB field code “22192”) to facilitate comparison with other datasets. Further details of the LTL measurements and their quality control has been reported elsewhere.^7^

For this analysis, from participants with LTL measurements in UKB, we excluded genetically related samples, randomly excluding one from each pair based on a kinship coefficient of K>0·088 (n=33,728) and samples with no genetic data or those that failed quality control (n=908). We also excluded participants who lacked information on ethnicity or white blood cell (WBC) count, which are both associated with LTL^7^ and were used together with age and sex to adjust the trait associations. This left a maximum of 422,797 participants available for the analysis (**Supplementary Figure 1**).

### Modifiable traits selection

We considered characteristics recorded at baseline in at least 50,000 UKB participants and identified 117 potentially modifiable traits, including three derived traits: waist–to–hip ratio, pulse pressure, and alcohol intake (**Supplementary Table 1**). These traits were grouped under 12 major categories, representing (alphabetically): alcohol intake (4 traits), anthropometry (12), blood biochemistry (13), cardiovascular (6), chronobiology (5), dietary intake (34), early life and sexual health (5), general health (11), physical activity (5), psychosocial (13), smoking (2) and socio–economic status (7) (**Supplementary Table 1**). While the extent to which some of the traits are modifiable can be debated, we chose to be inclusive in our selection, in order to evaluate as comprehensively as possible traits that could potentially influence LTL.

### Healthy behaviour indices

We generated our primary healthy behaviour index (HBI) based on a previously reported method.^19^ Healthy behaviour was considered as: 1) the absence of current or previous smoking; 2) engagement with physical activities that require the expenditure of more than 735 metabolic equivalents per hour in a week (METs/week); 3) maintenance of a healthy body weight assessed with a BMI in the range of 18·5 to 24·9 kg/m^2^; 4) adherence to a healthy diet and 5) moderate alcohol consumption (**Supplementary Table 2**). Adherence to a healthy diet, is characterised by a higher consumption of fruits, nuts, vegetables, whole grains, fish, and dairy products and a lower consumption of refined grains, processed meats, unprocessed red meats and sugar–sweetened beverages.^21^ A dietary index was created from the cumulative sum of the level of consumption of these components (theoretical range: zero to seven). Further details are provided in the **Supplementary Materials**. Participants with scores greater than or equal to 4 in the diet index were considered to follow a healthy diet. For the assessment of alcohol consumption, we considered participants’ self–reported weekly and monthly intake of different types of alcoholic beverages and converted this into grams of alcohol per day (see **Supplementary Materials**). Moderate alcohol intake was then considered as 5–15g of alcohol per day for women and 5–30g per day for men, respectively equivalent to ≤1 and ≤2 small wine glasses of 12% alcohol by volume.^19^ Alcohol intake for participants who reported current non– drinking (including formal drinkers) was assumed to be 0g of alcohol per day. For each behavioural component a score of one was given if the participant met the criterion for healthy behaviour, or zero otherwise. The scores were then summed to create our primary HBI that ranged from zero (no healthy behaviours) to five (adherence to all five healthy behaviours). There were 329,907 participants with complete information to calculate the primary HBI (**Supplementary Figure 1**).

To test the robustness of any association of LTL with healthy behaviour, we also created a second HBI based on another index that has been previously utilised.^20^ In this index, healthy behaviours were considered to be: 1) an absence of current smoking; 2) moderate or vigorous levels of physical activity; 3) maintenance of a healthy body weight assessed as a BMI less than 30 kg/m^2^ and 4) adherence to a healthy diet, while alcohol intake was not included (**Supplementary Table 2**). Participants were assigned a score of one if they met the criteria of a healthy behaviour component. The sum of these four scores provided a range from zero (no healthy behaviours) to four (adhering to all four healthy behaviour components). For this secondary HBI there were 331,658 participants with full information (**Supplementary Figure 1**). A comparison of the two HBIs and detailed scoring for each is shown in **Supplementary Table 2**.

### Statistical analysis

We undertook our main analysis on available data for each individual trait and each HBI. In addition, to assess the impact of missing trait values, particularly for the multivariable analysis, we repeated our analyses using multiple imputation (MI) by chained equations (MICE)^22^ with 10 imputed datasets. The imputation models included all modifiable traits, medical conditions, age, sex, ethnicity and WBC and we specified linear, logistic and multinomial regression imputation models for continuous, binary and categorical traits respectively. To ensure convergence, we performed 10 iterations for each imputed dataset.^22^ We compared the Monte Carlo error against the standard errors (SEs) of the estimated betas (parameters) of the various traits to evaluate the performance of the imputation.

#### Single traits

To identify traits significantly associated with LTL we fitted a linear regression model for each trait with z–standardised LTL as the response variable, adjusting for age, sex, ethnicity and WBC count (as these variables have been shown to be associated with LTL^7^). In these analyses, continuous traits were first winsorized at the 0·5% and 99·5% percentile values to exclude extreme outliers. Traits were loge–transformed where appropriate after graphically checking their distributions, before being scaled to the standardised normal distribution to aid interpretation. For continuous traits results are shown as beta coefficients per 1 SD increase in the trait, while for binary traits results are presented as beta coefficients from a comparison of yes to no status. Statistical significance for the trait analyses was set at a Bonferroni corrected threshold of P<4·27×10^-4^ to account for the number of traits analysed. To aid interpretation, we calculated the association with each trait in terms of number of equivalent years of age–related change in LTL, by dividing the beta coefficient for the trait by the absolute value of the beta coefficient for the age–related change in LTL (-0·023 per year).^7^ Because of potential correlation between traits associated with LTL, to identify those that had the strongest independent associations with LTL, we also fitted a multivariable linear regression model restricted to traits that were Bonferroni significant and had an association with LTL equivalent to an age effect on LTL of ≥2 years in the analysis on the available data.

#### Healthy behaviour indices

To examine the association between the HBIs and LTL, we also used linear regression models. In the base model, we analysed the association between the HBI and LTL adjusting for age, sex, ethnicity and WBC count. As behaviours may be affected by the presence of disease we next adjusted for presence of several self–reported doctor diagnosed chronic medical conditions including diabetes (UKB field code “2443”), cancer (“2453”), vascular disease and hypertension (both derived from field “6150”) at the time of enrolment into UKB (adjusted model). Finally, to account for potential confounding by other modifiable traits which were not directly considered as part of the HBIs, we additionally adjusted for the most significant trait(s) as proxies for each category, presented in **Supplementary Table 1** (final model). Specifically, we adjusted for higher educational level (as proxy for socio–economic status), insomnia (as proxy for chronobiology), fed–up feelings (as proxy for psychosocial well–being) and low–density lipoprotein (LDL)– cholesterol, C–reactive protein and estimated glomerular filtration rate (eGFR) as representatives of biochemical traits.

#### Mendelian randomisation analysis

Two of the traits that showed the strongest association with LTL in both the single and multivariable analysis were educational attainment and current smoking. To investigate the causality and directionality of these associations, we undertook bi–directional Mendelian Randomisation (MR) analysis, using large–scale genome–wide association study (GWAS) datasets, for LTL, education attainment and smoking behaviour.^4,23,24^ For educational attainment, to assess if the direction of effect was from education to LTL, we used genetic variants that were associated with number of years of schooling (EduYears) in a GWAS of 1·1 million individuals^23^ as MR instruments and examined their association with LTL in the LTL GWAS dataset.^4^ For smoking, we used genetic variants associated with two smoking phenotypes: initiation of regular smoking (a binary phenotype indicating whether an individual had ever smoked regularly (current or former) or not, and smoking intensity (cigarettes smoked per day for both current and former smokers).^24^ In each case, to examine the converse possibility, i.e. that the direction of association is from LTL to educational attainment and smoking, we used genetic variants that we have recently reported to be associated with LTL in UK Biobank,^4^ and studied their associations with educational attainment and smoking behaviour in the respective GWASs. Further details of the MR analysis and the number of genetic variants used in each analysis are provided in the **Supplementary Material**.

#### Clinical outcomes

To evaluate the potential clinical relevance of our findings, we undertook two types of analyses. First, we examined whether the association of LTL with various diseases, where we have previously found evidence of a potential causal association in UKB through MR analysis,^4^ differs according to the number of healthy behaviours, using the primary HBI for this analysis. Second, as we have previously shown^4^ that longer LTL is associated with greater life expectancy and lower risk of CAD, and healthy behaviours also impact on these phenotypes, we calculated the proportion of any association of primary HBI on life expectancy and risk of CAD that may be mediated through LTL. Further details of these analyses are provided in the **Supplementary Material**.

## Results

### Demographic characteristics

Characteristics of the 422,797 UKB participants studied are summarised in **Table 1**. The participants were aged 40–69 at recruitment (mean 56·6 (SD 8·0) years) with more women (53·8%) and predominantly of white ethnicity (94·6%).

**Table 1:**
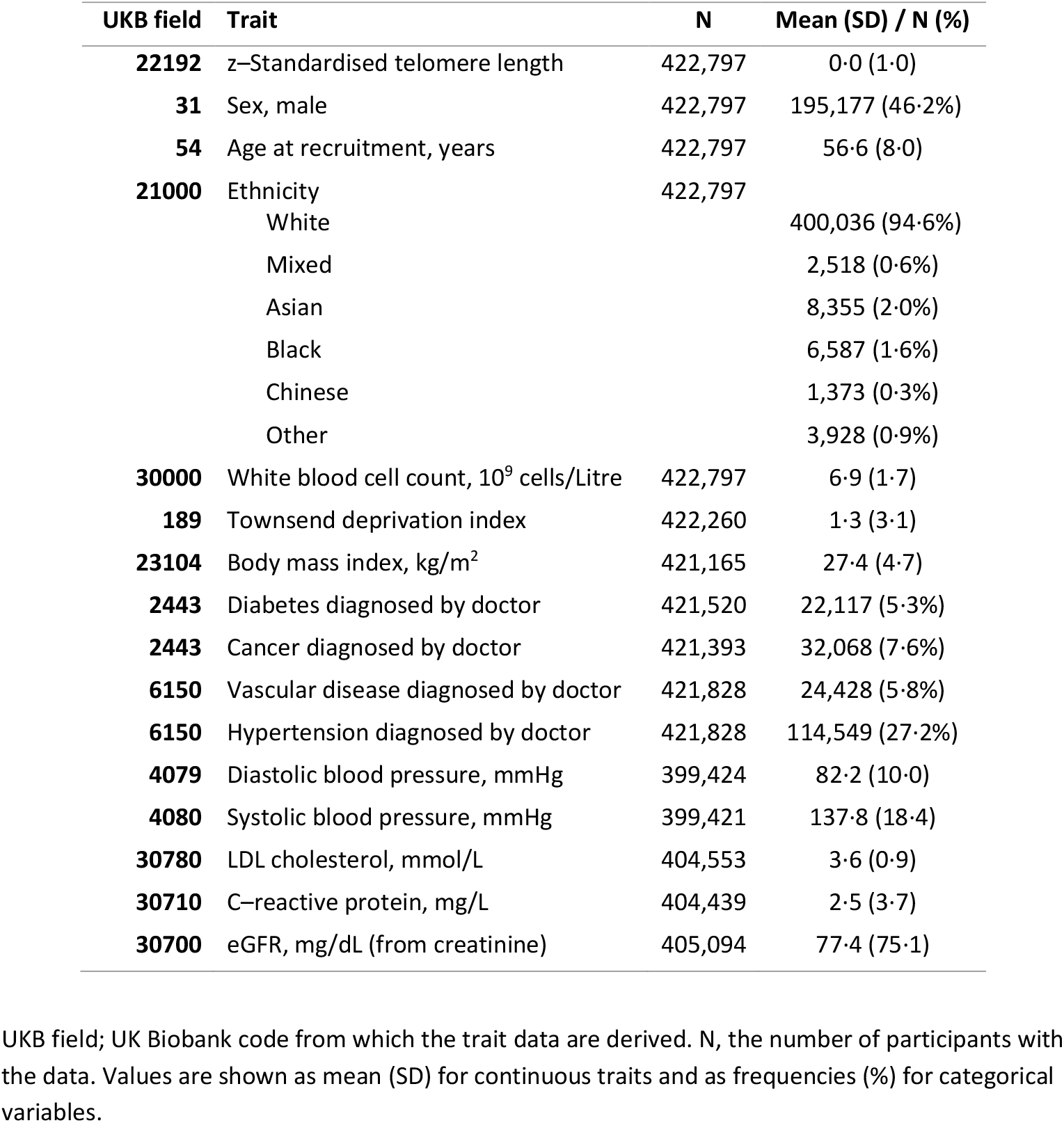
Selected characteristics of the UK Biobank participants analysed.

| UKB field | Trait | N | Mean (SD) / N (%) |
| --- | --- | --- | --- |
| **22192** | z-Standardised telomere length | 422,797 | 0.0 (1.0) |
| **31** | Sex, male | 422,797 | 195,177 (46.2%) |
| **54** | Age at recruitment, years | 422,797 | 56.6 (8.0) |
| **21000** | Ethnicity | 422,797 |  |
|  | White |  | 400,036 (94.6%) |
|  | Mixed |  | 2,518 (0.6%) |
|  | Asian |  | 8,355 (2.0%) |
|  | Black |  | 6,587 (1.6%) |
|  | Chinese |  | 1,373 (0.3%) |
|  | Other |  | 3,928 (0.9%) |
| **30000** | White blood cell count, 10<sup>9</sup> cells/Litre | 422,797 | 6.9 (1.7) |
| **189** | Townsend deprivation index | 422,260 | 1.3 (3.1) |
| **23104** | Body mass index, kg/m<sup>2</sup> | 421,165 | 27.4 (4.7) |
| **2443** | Diabetes diagnosed by doctor | 421,520 | 22,117 (5.3%) |
| **2443** | Cancer diagnosed by doctor | 421,393 | 32,068 (7.6%) |
| **6150** | Vascular disease diagnosed by doctor | 421,828 | 24,428 (5.8%) |
| **6150** | Hypertension diagnosed by doctor | 421,828 | 114,549 (27.2%) |
| **4079** | Diastolic blood pressure, mmHg | 399,424 | 82.2 (10.0) |
| **4080** | Systolic blood pressure, mmHg | 399,421 | 137.8 (18.4) |
| **30780** | LDL cholesterol, mmol/L | 404,553 | 3.6 (0.9) |
| **30710** | C-reactive protein, mg/L | 404,439 | 2.5 (3.7) |
| **30700** | eGFR, mg/dL (from creatinine) | 405,094 | 77.4 (75.1) |

UKB field; UK Biobank code from which the trait data are derived. N, the number of participants with the data. Values are shown as mean (SD) for continuous traits and as frequencies (%) for categorical variables.

### Single traits and LTL

Of the 117 potentially modifiable traits analysed, 71 (60·7%) showed a significant association with LTL after adjusting for multiple testing (P<4·27×10^-4^) (**Figure 1, Supplementary Table 1 and Supplementary Figure 2**). The findings from the multiple imputation analysis were highly concordant (**Supplementary Table 1**). For many of the traits, although the associations were significant, the beta coefficients were small and equivalent to less than one year of age–related change in LTL (**Supplementary Table 1**). However, for 17 traits the associations were more substantial (≥ than 2 years of age–related change in LTL) per 1 SD change in the trait or group contrast (**Figure 2**). As some of these traits may be correlated with each other, a multivariable regression analysis was undertaken to identify those with the strongest independent associations. From the available data, this was only possible in 84,462 participants with data on all 17 traits. The univariate associations for these traits in this subset of participants were similar to those seen in the fully available data for each trait (**Supplementary Table 3**). In the multivariable analysis, only three traits retained a significant association equivalent to ≥2 years age–related change in LTL (**Supplementary Table 3**). Compared those who had no intake those who had an intake of oily fish of 2–4 portions a week had longer LTL equivalent to 2·4 years of age–related change in LTL (P=5·5×10^-5^); compared with those who reported excellent overall health status, those with poor overall health status on average had shorter LTL equivalent to 4·0 years of age–related change in LTL (P=4·1×10^-4^); and compared to those with education below end of secondary school those who attained degree level education had longer LTL equivalent to 2·8 years of age–related change in LTL years (P=1·6×10^-6^). For all three traits we saw a gradient of difference in LTL across more granular levels of the trait adding confidence to the findings (**Supplementary Table 3**). In the multivariable multiple imputation analysis for these 17 traits, both intake of oily fish and educational attainment showed very similar associations as seen with the available data (**Supplementary Table 3**). The magnitude of the association with overall health status became less strong (equivalent to 2·0 years of age–related change in LTL) but remained significant (P=4·2×10^-7^), while the associations with brisk walking pace with longer LTL (equivalent to 2·2 years of age–related change in LTL, P=3·3×10^-14^) and current smoking with shorter LTL (equivalent to 2·0 years of age–related change in LTL, P=3·5×10^-17^) became stronger (**Supplementary Table 3**).

**Figure 1.**
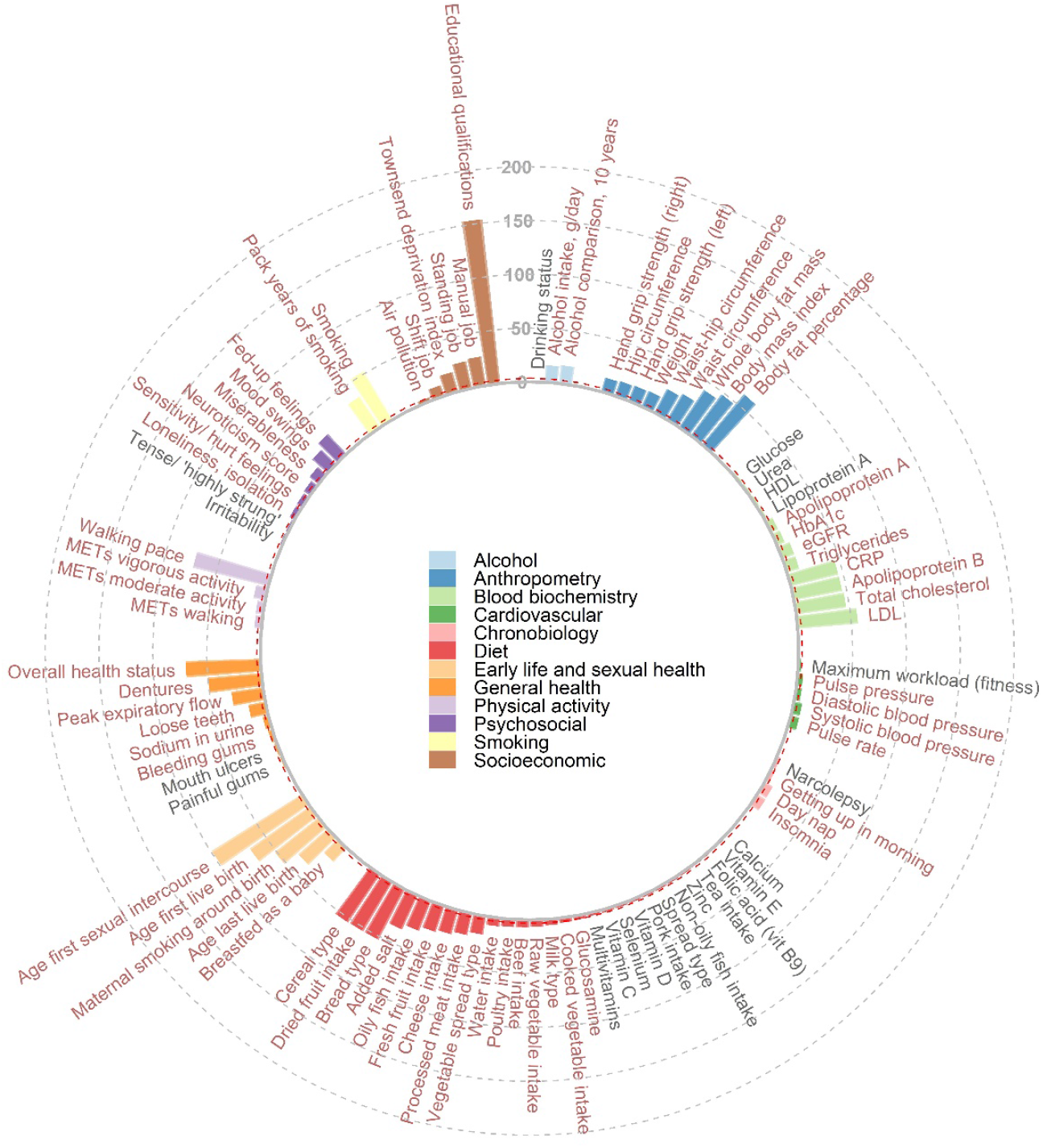
A circular plot showing traits nominally associated with LTL. For each trait the P–value shown is from a univariate linear regression model adjusted for age, sex, ethnicity and white blood cell count. Traits are grouped and coloured by category with –log10 (P–value) on the Y–axis, indicated by the axis labels at 12 o’clock. For categorical traits the global P–value from a likelihood ratio test is shown. Bonferroni significant traits (P<4·27×10^-4^) have red text, nominally significant trait (P<0·05) have black text. Non–significant traits (P>0·05) are not shown in the figure (these are shown in Supplementary Table 1).

**Figure 2.**
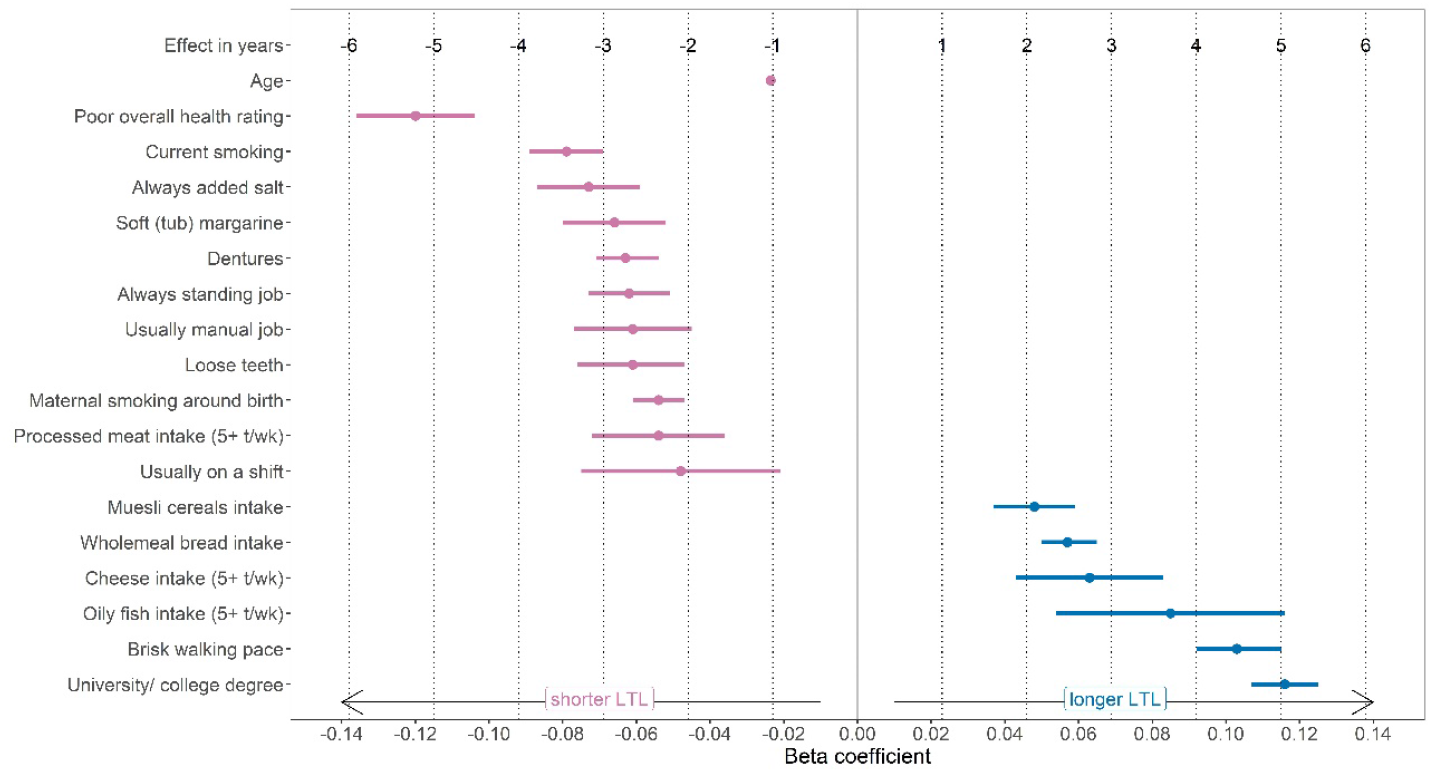
Individual traits most strongly associated with LTL. The traits are shown on the Y–axis and the beta coefficients for the associations with LTL on the X–axis. Effect in years is the ratio of the trait beta and the absolute value of the age beta (-0·023). Continuous traits are estimated for a 1 SD increase in the trait. Binary traits compare yes to no. Poor overall health rating is compared to Excellent. Current smoking is compared to Never. Always added salt is compared to Never/rarely. Soft (tub) margarine compared to Olive–oil spread. Always standing job/Usually manual job are compared to Never/rarely. Processed meat/Cheese/Oily fish intake (5+ t/wk) are compared to Never. Usually on a shift is compared to Never. Muesli cereals compared to Other types. Wholemeal bread intake is compared to White bread. Brisk walking pace is compared to Slow. University/college degree is compared to No qualifications.

### Health behaviour indices and LTL

For the primary HBI, 4·4% (14,576) of participants had a score for zero for healthy behaviour, 20·9% (69,082) had a score of one, 34·6% (114,086) had a score of two, 27·6% (90,985) had a score of three, 10·8% (35,752) had a score of four, and 1·6% (5,436) had a score of five. The characteristics of participants partitioned by primary HBI score is shown in **Table 2**. Although statistically significant, there was no substantial difference in the mean age of participants across the scores and there was only minor variation in proportion of women (**Table 2**). However, several social and clinical characteristics showed a substantial gradient in prevalence from those with no healthy behaviours (score zero) to those with a full set of healthy behaviours (score five). This was further reflected in the prevalence of several diseases. For example, the prevalence of diabetes was 11·1% among those with a score of zero compared with 1·5% in those with a score of five. Likewise, the prevalence of hypertension and cardiovascular disease also showed around a 3–fold difference between these groups (**Table 2**).

**Table 2:**
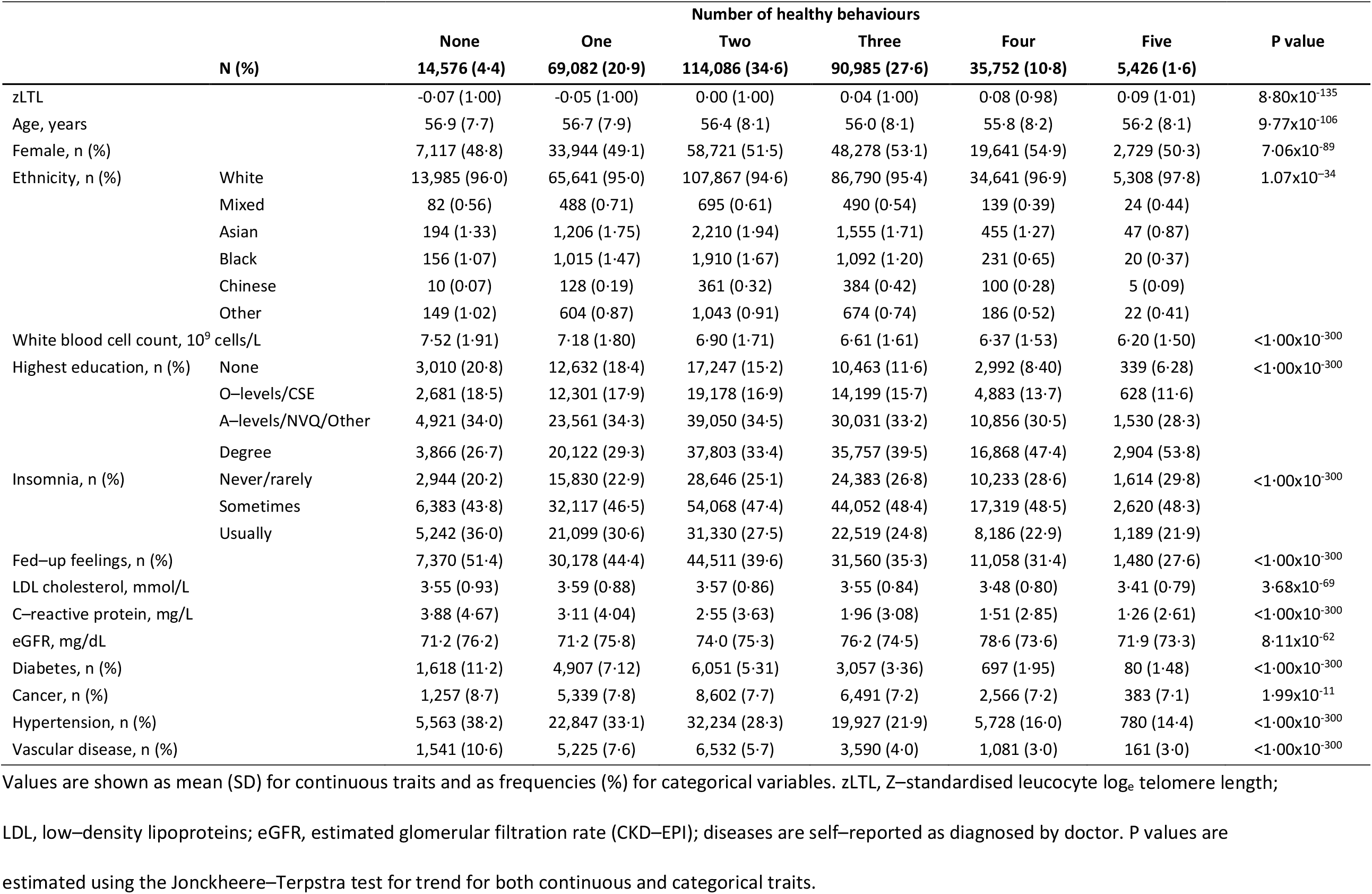
Participant demographics partitioned by number of healthy behaviours for the primary HBI (n= 329,907).

For the primary HBI, in the base model (adjusted for age, sex, ethnicity and WBC count) those with adherence to all healthy behaviours (score five) had significantly longer LTL on average (beta coefficient=0·107, 95% CI: 0·077–0·137, P=5·1×10^-12^), compared to those with no healthy behaviours (score zero) (**Supplementary Table 4**). This is equivalent to 4·6 years in age–related change in LTL. Adjusting the model for the presence of chronic health conditions did not alter the association (beta coefficient=0·103, 95% CI: 0·072–0·133, P=3.9×10^-11^) (**Supplementary Table 4**). The association was modestly attenuated after further adjustments for higher educational level, insomnia, fed–up feelings, LDL–cholesterol, C–reactive protein and eGFR, but remained significant (beta coefficient=0·072; 95% CI: 0·041–0·104, P=7·7×10^-06^), equivalent to 3·1 years of age–related change in LTL (final model, **Supplementary Table 4**). Across the primary HBI score, there was a consistently additive dose–response association of greater healthy behaviours with longer LTL (**Figure 3**). The findings were very similar from the multiple imputation analysis across all models (**Supplementary Table 4**). For example, in the final model the beta coefficient was 0·074 for the imputed data (**Supplementary Table 4**). Overall, the primary HBI explained 0·180% (0·178%–0·181%) of the variation in LTL.

**Figure 3.**
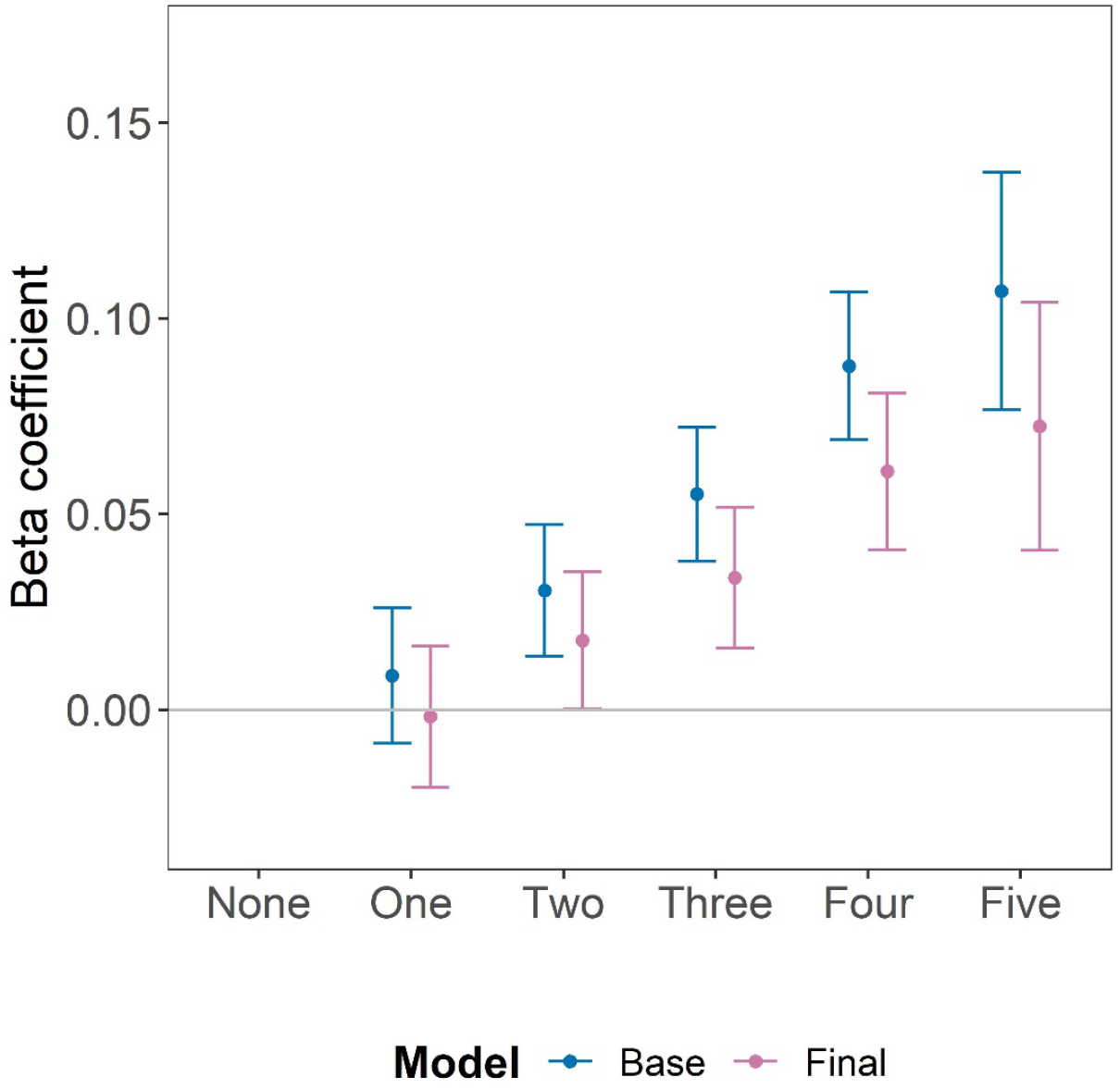
Association between the number of components of the primary healthy behaviour index and LTL. In the Base model adjustments were made for age, sex, ethnicity and white blood cell count and in the Final model additional adjustments were made for self–reported diagnosed by doctor of chronic medical conditions (diabetes, cancer, hypertension and vascular disease), insomnia, fed–up feelings, LDL–cholesterol, C–reactive protein, eGFR, and educational attainment. Bars represent the 95% confidence intervals around the beta coefficient.

Despite the composition of the second HBI being different (**Supplementary Table 2**) and the proportion of participants across the scores being very different to that for the primary HBI (**Supplementary Table 5**), we found the same pattern of positive linear association of greater adherence to healthy behaviours with longer LTL (**Supplementary Table 6**). In the base model for the second HBI in the available data, the beta coefficient was 0·117 (95% CI: 0·074–0·160, P=1·1×10^-07^) for those with a score of four compared to a score of zero, equivalent to 5·1 years of age–related change in LTL and in the fully adjusted model it was 0·084 (95% CI: 0·039–0·129, P=2·9×10^-04^), equivalent to 3·7 years of age–related change in LTL, with again highly concordant findings in the fully imputed data (**Supplementary Table 6**).

#### Mendelian randomisation analysis

In the MR analysis, there was a positive association between number of years spent in education and LTL. One year increase in number of genetically–determined years spent in education was associated with longer LTL equivalent to 4·2 years (95% CI: 3·5–4·9 years) of age–related change in LTL (P=3·19×10^-30^) (**Supplementary Table 7a**). There was no evidence of pleiotropy (MR Egger P=0·15) and other MR methods supported the findings (**Supplementary Table 7a**). The inverse MR analysis showed no evidence that LTL is causally associated with number of years spent in education (**Supplementary Table 7a**). Similarly, MR analysis supported a casual association of smoking behaviour with LTL with no evidence of reverse causation (**Supplementary Tables 7b and 7c**). A history of genetically–determined smoking (current or former) was associated with shorter LTL equivalent to 2·5 years (95% CI: 1·7–3·2) of age–related change in LTL (P=1·4×10^-10^) (**Supplementary Tables 7b)** while 1 SD genetically–determined increase in cigarettes smoked per day was associated with shorter LTL equivalent to 2·8years (95% CI: 1·1–4·5) of age–related change in LTL (P=0·003) (**Supplementary Tables 7b and 7c)**.

### Clinical consequences

Out of 22 disorders for which we had previously seen concordant genetic and observational association with LTL, for only one disease, sarcoma, did we see a possible interaction between HBI and LTL, with those with 4–5 healthy behaviours having an inverse association between LTL and risk of sarcoma while those with lower scores for healthy behaviours had a positive association (P=0·002) (**Supplementary Table 8 and Supplementary Figure 3**). However, when accounting for the number of tests performed, this was not significant (Bonferroni P value=1·14×10^-3^) and there were only 24 incident cases among those with 4–5 healthy behaviours, necessitating caution in interpreting this finding.

A higher HBI score was associated with longer life expectancy in both men and women throughout the age range from 40 to over 85 years (**Supplementary Figure 4**). At age 40 years, men with a primary HBI score of five had on average 10·4 (95% CI: 7·5–13·3) years longer life expectancy compared to men with a HBI score of zero. Corresponding estimates in women were 9·4 (95% CI: 6·2–12·6) years longer life expectancy. Neither of these associations were significantly changed by adjustment for LTL (**Supplementary Figure 4**). Each unit higher primary HBI was associated with a 21% (95% CI: 20%–22%) lower risk of CAD. The proportion of this risk that was mediated through LTL, although significant was small (0·49% (95% CI: 0·33%–0·65%, P=2·8×10^-09^) (**Supplementary Figure 5**). The findings were similar for the second HBI (**Supplementary Figure 5**).

## Discussion

In the last two decades compelling evidence has emerged that variation in telomere length affects multiple physiological traits as well as risk of several diseases. This has raised two important and hitherto unresolved questions: first, the extent to which telomere length may be modifiable through lifestyle factors and specific behaviours and second, whether any such effect has important clinical consequences. Here, by generating LTL measurements at large scale in UK Biobank and utilising the extensive information on lifestyle, behaviour and disease outcomes that have been collected on participants we provide important new insights into these two questions.

In single trait analysis of 117 potentially modifiable traits, we found demonstrable associations of LTL with many traits. Quantitatively, the associations were generally small and equivalent to less than one year of age–associated change in LTL. For example, consistent with previous reports,^11,13^ we observed a small inverse association between BMI and LTL (**Supplementary Table 1**). However, 17 traits showed stronger association equivalent to ≥2 years of age–related change in LTL (**Figure 2**). In a multivariable analysis of these traits in the subset of participants in whom data were available on all of these traits, three remained significant and retained the larger effect sizes: oily fish intake, educational attainment and general health status. Similar associations for these traits were seen in the multiple imputation analysis which additionally also showed significant associations with brisk walking pace and with current smoking (**Supplementary Table 3**).

The association of higher oily fish intake with longer LTL is interesting because a previous longitudinal study showed that higher intake of marine omega–3 fatty acids was independently associated with a lower rate of telomere shortening over 5 years^25^ suggesting a possible causal association. Using MR analysis, we have recently reported that the association of brisk walking pace with longer LTL is also likely to be causal.^26^ Additionally, here our MR analysis of educational attainment and smoking behaviour suggests that the associations of these traits with LTL are also likely to be causal. Although the exact phenotypes used to identify the genetic variants in the GWAS analyses of education (number of years in schooling) and smoking behaviours (smoking initiation and smoking intensity) are not identical to the phenotypes available for these traits in UKB and analysed here (educational attainment and current smoking), in both instances the different phenotypes are likely to be highly correlated. For smoking, the finding that both genetically–determined smoking initiation and higher smoking intensity both were associated with LTL provides further support for a causal association. Furthermore, we found no genetic evidence that LTL affects the duration of time in education or smoking behaviour to support the alternate explanation for the observed association between these traits and LTL. In aggregate, our findings indicate that at least some of the observed associations between the traits studied and LTL are likely to be causal.

While the associations of individual traits with LTL are interesting perhaps the more important question from a clinical perspective is whether particular forms of healthy behaviour are associated with LTL, either individually or in aggregate. We calculated two previous reported indices of such behaviours each comprising evaluation of smoking, physical activity, diet and maintenance of a healthy body weight and in one index, alcohol intake. While the criteria for each behaviour in the two indices partitioned participants differently, we saw a similar pattern of relationship with LTL with both indices. In each case, the healthy behaviour score was positively and linearly associated with longer LTL. The finding was consistent between the analysis on participants with available data and the analysis on all participants after imputation of missing trait values. The association was not attenuated by the presence of several common chronic conditions (cancer, diabetes, hypertension and vascular diseases) which might have altered behaviours and was only modestly attenuated by adjustment for other factors not included in the indices that differed between healthy behaviour groups. Notably, although CRP levels varied between the HBI groups, being higher in those with fewer healthy behaviours, adjustment for it did not remove the association, suggesting that any effect on systemic inflammation may only explain part of the association between healthy behaviours and LTL.

Despite the reproducible association of healthy behaviours with LTL, the amount of inter–individual variation explained by such behaviours is very modest, accounting for less than 0·2% of such variation. To put this into context, age, sex, ethnicity and WBC count together explain about 5·5% of variance in LTL.^7^ LTL is largely genetically determined, with heritability estimates of ∼0·70.^5^ The low variance explained by health behaviours probably explains why we did not see a significant influence of such behaviours on the association between LTL and various diseases. Similarly, although a higher HBI score and longer LTL are both associated with greater life expectancy and lower risk of CAD,^4^ we found no evidence that a substantial proportion of the association of HBI with either is mediated through LTL.

Our findings have several individual and population–level implications. They demonstrate that telomere length may be modifiable by lifestyle and behaviour. However, the quantitative effects of these are small compared to the much larger genetically–driven variation in LTL between individuals. As such, attempts to modify the association between LTL and risk of various diseases through adoption of healthy behaviours or targeting a particular modifiable trait are likely to have modest effects at best. Instead, the focus should be on pharmacological^27^ methods to modify telomere length. Alternately, developing a better understanding of the mechanisms by which variation in telomere length affects risk of diseases and then selectively targeting these mechanisms may be fruitful. For example, there is evidence that variation in telomere length affects the development of clonal haematopoiesis of indeterminate potential (CHIP) which in turn is associated with risk of CAD.^28^ Therefore, targeting CHIP could, at least partly, attenuate the association of LTL with risk of CAD.

Strengths of our study include its large scale and the detailed phenotyping available in UK Biobank to enable a comprehensive and robust analysis of the association between modifiable traits and behaviours and LTL. However, as with any cross–sectional analysis caution is necessary about making causal inferences, despite supportive genetic analysis. Furthermore, because of a lack of longitudinal telomere measurements, we are unable to assess whether within–individual changes in LTL with age are affected by modifiable traits and behaviours and whether these have clinical consequences. Finally, UK Biobank is under–represented in non–white ethnicities and there is evidence that the cohort recruited participants that were healthier than the general population.^29^ It is therefore possible that the magnitude of the associations of modifiable traits and healthy behaviours with LTL may vary in other populations and further studies are necessary.

In summary, we show that several potentially modifiable traits as well as healthy behaviours have a quantifiable association with LTL. At least in some cases this association is likely to be causal. However, the effect on LTL is modest and while modifying these traits and behaviours may have other benefits in their own right, they are unlikely to substantially alter the association between LTL and the risk of major disease outcomes.

## Data Availability

The MTA precludes direct data sharing. The LTL measurements have been deposited back to the UK Biobank and can be obtained by application to UKB.

## Contributors

VB, CN, EA, CPN, VC and NJS developed the analysis plan and curated the list of modifiable traits; VB and CM conducted the majority of the analysis including creation of the healthy behaviour indices; QW provided input into the MR analysis; SK carried out the life expectancy analysis; AMW provided advice and oversaw the multiple imputations; EDI, ASB, JRT, JND and AMW reviewed findings and provided important feedback on additional analyses; VB, CM, CPN, VC and NJS prepared the manuscript and all authors provided critical revision. VC, CPN, JRT, JND and NJS (Principal investigator) secured funding and oversaw the generation and curation of the LTL measurements. VPN, CM, CPN, VC and NJS verified the underlying data. All authors had full access to the data and accept responsibility to submit for publication.

## Declaration of interests

The authors have no conflicts or interests to declare.

## Data Sharing

The LTL measurements have been deposited in UKB. These measurements as well as other data used in this paper can be obtained by application to UKB.

## Acknowledgements

This work was funded by the UK Medical Research Council (MRC), Biotechnology and Biological Sciences Research Council and British Heart Foundation (BHF) through MRC grant MR/M012816/1. CPN is funded by the BHF (SP/16/4/32697). VC, CM, VB, QW, CPN and NJS are supported by the National Institute for Health Research (NIHR) Leicester Cardiovascular Biomedical Research Centre (BRC–1215–20010). Cambridge University investigators are supported by the BHF (RG/13/13/30194; RG/18/13/33946), Health Data Research UK, NIHR Cambridge Biomedical Research Centre (BRC–1215–20014), NIHR Blood and Transplant Research Unit in Donor Health and Genomics (NIHR BTRU–2014–10024) and MRC (MR/L003120/1). JD holds a BHF Personal Professorship and NIHR Senior Investigator Award. AMW and EA received support from the EU/EFPIA Innovative Medicines Initiative Joint Undertaking BigData@Heart (11607).

## Notes

### Competing Interest Statement

The authors have declared no competing interest.

### Author Declarations

UKB received approval from the North West Centre for Research Ethics Committee (11/NW/0382) and have therefore been performed in accordance with the ethical standards laid down in the 1964 Declaration of Helsinki and its later amendments. All participants gave written informed consent to participation. The use of data presented in this paper was approved by the Access Committee of UKB under application number 6077.

