## Supplementary materials for "Modifiable traits, healthy behaviours, and leucocyte telomere length"

Vasiliki Bountziouka et al.

**Index**

**A. Supplementary Methods**

1. Assessment of adherence to healthy diet Page 3

2. Bi-directional Mendelian Randomisation analysis Page 4

3. Effect of healthy behaviours on the association of LTL with diseases Page 4

4. Mediation analysis Page 5

5. Supplementary references Page 6

**B. Supplementary Figures**

**Supplementary Figure 1.** Flowchart of participants included in different analyses and reasons for exclusion. Page 8

**Supplementary Figure 2.** Number of modifiable traits associated with LTL by categories and significance level. Page 9

**Supplementary Figure 3.** Association of LTL with risk of 22 diseases across different scores for the primary healthy behaviour index. Page 10

**Supplementary Figure 4.** Years of life gained according to healthy behaviour index (HBI) groups with/without adjustment for leucocyte telomere length (LTL) as a mediator. Page 11

**Supplementary Figure 5.** Mediation analysis to examine the proportion of the association of the healthy behaviour indices with CAD that may be mediated through LTL. Page 12

**C. Supplementary Tables**

**Supplementary Table 1:** Univariate model results for the association of modifiable traits with LTL. Page 13

**Supplementary Table 2:** Scoring system for the primary and second healthy behaviour indices. Page 20

**Supplementary Table 3:** Multivariable model results for 17 traits with an association with LTL equivalent to ≥2 years of age–related change in LTL. Page 21

**Supplementary Table 4:** Multivariable model results for the association of primary healthy behaviour index with LTL.

Page 24

**Supplementary Table 5:** Participant demographics by healthy behaviour groups of the second healthy behaviour index. Page 25

**Supplementary Table 6:** Multivariable model results for the association of second healthy behaviour index with LTL. Page 27

**Supplementary Table 7:** Bi–directional Mendelian Randomisation analysis. Page 28

**Supplementary Table 8:** Multivariate model results for the risk of selected diseases per SD longer of LTL, overall and stratified by number of healthy behaviours. Page 30

**Supplementary methods**

**1. Assessment of adherence to healthy diet**

Adherence to a healthy diet, is characterised by the increased consumption of fruits, nuts, vegetables, whole grains, fish, and dairy products and a reduced consumption of refined grains, processed meats, unprocessed red meats and sugar-sweetened beverages.[1] Similarly to an established rationale,[2] we identified six main food groups (vegetables (UKB field codes "1289" and "1299"), fruit ("1309" and "1319"), fish ("1329" and "1339"), type and number of slices/ bowls of bread ("1438" and "1448") and cereals ("1458" and "1468"), red meat intake ("1369", "1379", and "1389") and processed meat ("1349")), obtained via a touchscreen food frequency questionnaire, and we created an index to measure healthy diet as follows. A score of one was assigned if participants reported increased consumption of fruit (≥3 portions/day), vegetables (≥3 portions/day), fish (daily/ weekly consumption) or decreased consumption of processed meat (consumed never/ rarely) and other red meats (from never to monthly intake). A score of two was assigned if participants reported increased consumption of whole grain bread and cereals (≥3 portions/day), whilst a score of one was assigned if they reported decreased consumption of white bread and refined cereals (≤1·5 portions/day). A portion was considered to be four heaped tablespoons of vegetables, one medium-sized piece of fruit, two slices of bread and one bowl of cereals.[3] In all food groups a score of zero was assigned otherwise. A dietary index was created as the cumulative sum of these six components (theoretical range: zero to seven). Participants with scores greater than or equal to four in the diet index were considered to follow a healthy diet.

For the assessment of moderate alcohol consumption we considered participants' self–reported weekly and monthly intake in terms of glasses of red wine (UKB field codes "1568" and "4407" respectively), champagne/ white wine ("1578", "4418"), beer/ cider ("1588", "4429"), spirits ("1598", "4440"), fortified wine intake ("1608", "4451"), and other alcoholic drinks ("5364", "4462") and converted it into average daily intake by dividing by 7 or 30 accordingly. Number of drinks per day were quantified as the number of UK units of alcohol intake and then converted to grams of alcohol (1 unit=8g of alcohol).[4] Moderate alcohol intake was then considered as 5–15g of alcohol per day for women and 5–30g per day for men.[5] Alcohol intake for participants who self–reported as non–drinking were assumed to intake 0g of alcohol per day.

**2. Bi-directional Mendelian Randomisation analysis**

To investigate the directionality of the observed associations between educational attainment and smoking behaviour and LTL, we undertook bi–directional mendelian randomisation (MR) analysis, using large–scale genome–wide association study (GWAS) datasets[6,7,8] for the three traits. To examine if educational attainment was causal for a change in LTL, of the 1,271 independent variants associated with number of years spend in education (EduYears) in the educational attainment GWAS[6] we were able to match 1,267 SNP in the LTL GWAS.[8] For the reverse MR analysis (LTL to EduYears) we were able to match 87 out of 130 LTL–associated genetic variants in the educational attainment GWAS. For the smoking phenotypes we able to match 374 of the 378 independently associated variants for smoking initiation and all 55 for smoking intensity in the LTL GWAS data. Of the 130 variants for LTL we were able to match 89 in both the smoking initiation and smoking intensity GWAS data.

For each analysis, we used the inverse–variance weighted MR method[9] allowing for a random effect to estimate the causal association and also reported the P–value for the intercept from MR Egger[10] as a check for pleiotropy. As sensitivity analyses, we undertook MR analyses using the Weighted Median method[11] which is additionally robust in the presence of outliers and the MR Raps method[12] which overcomes challenges related to measurement error, weak or invalid (due to pleiotropy) measurements and selection bias (due to weak instrument). Therefore a combination of these methods provides the best evidence for the presence of a causal association. All MR analyses were performed in R version 3·1·6.[13]

**3. Effect of healthy behaviours on the association of LTL with diseases.**

To investigate whether healthy behaviours affected the association of LTL with disease, we selected 22 diseases where we had previously seem evidence of a possible causal association at either at Bonferroni ($4·1x10^{-4}$; 12 diseases) or nominal ($5·0x10^{-2}$; 10 diseases) significance level.[8] For all diseases there was a concordant observational association with LTL.[8] To define incident cases for each disease, we used hospital episode statistics (HES) data using primary and secondary codes from the 9th and 10th revisions of the international statistical classification of diseases and related health problems (ICD–9 & 10) and the office of population censuses and surveys classification of surgical operations versions 3 and 4 (OPCS–3 & 4) from the UK office of national statistics, as previously described.[6] We used the date of sample collection that LTL was measured as the baseline date, and censored the end of follow–up in hospital health record data as 31st March 2020. Data about deaths was also subject to censoring using the same date of 31 March 2020 used for HES records. Cases that were self–reported at baseline or had a recorded hospitalisation with any (primary or secondary) diagnosis of the disease were excluded from the analysis. Incident cases were then defined as the first recorded event (primary or secondary) of the disease occurring after the UKB baseline visit. Time–to–event is defined as the post–baseline date of the first incident hospitalisation or death, or otherwise censored at the end of study follow–up on 31 March 2020.

To examine whether the association of LTL with incident diseases varied with the number of healthy behaviours we utilised a cox-regression model adjusted for a) age, sex, ethnicity and WBC (base model), b) additionally adjusted for previously diagnosed diabetes, cancer, hypertension, vascular disease (adjusted model) and c) further adjusted for educational level, insomnia, fed–up feelings, LDL–cholesterol, C–reactive protein, estimated glomerular filtration rate (CKD-EPI) (full model), also allowing for interactions between the covariates and the primary HBI as appropriate. A Wald test was used to decide on the overall significance of the interaction terms at the 5% level. Results are given as hazard ratios (95% confidence interval), with the point estimates being corrected for the regression dilution ratio of 0·68 for LTL measurements as described elsewhere.[14] As this analysis included two models for 22 diseases, the significance level was set at $1 \cdot 14 \times 10^{-3}$.

**4. Mediation analysis**

To assess the extent to which any association of healthy behaviours on life expectancy could be mediated through an effect on LTL, we first computed the association of the primary HBI score with life expectancy using public health modelling methods previously described[8] that combine cause–specific mortality rates from the general population and age–specific hazard ratios (HRs) for mortality to estimate differences in life expectancy for different primary HBI score groups using the group with a score of zero as reference. The extent of any mediation through LTL was then assessed by comparing differences in life expectancy estimated when applying HRs with/without adjustment for LTL.

We used structural equation models[15] to examine whether, and to what extent, the effect of the healthy behaviours on coronary artery disease (CAD) risk is mediated through LTL. In this analysis, the association of the primary HBI with risk of CAD are presented as a continuum i.e. per unit increase in the HB score. Results of the mediation analysis are shown as the percentage of the total effect of the primary HBI on CAD risk that could be mediated through LTL.

### Supplementary Figures

**Supplementary Figure 1. Flowchart of participants included in different analyses and reasons for exclusion.**

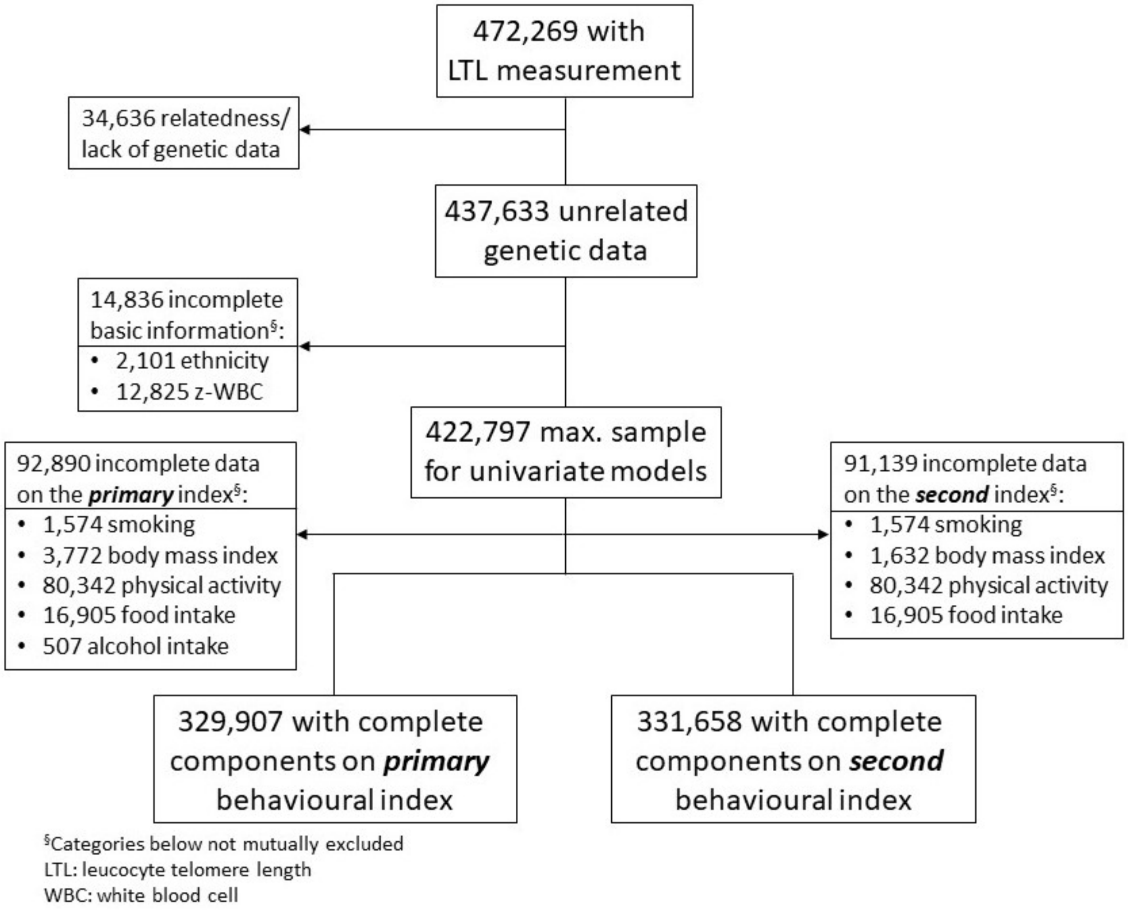

**Supplementary Figure 2. Number of modifiable traits associated with leucocyte telomere length by categories and significance level.**

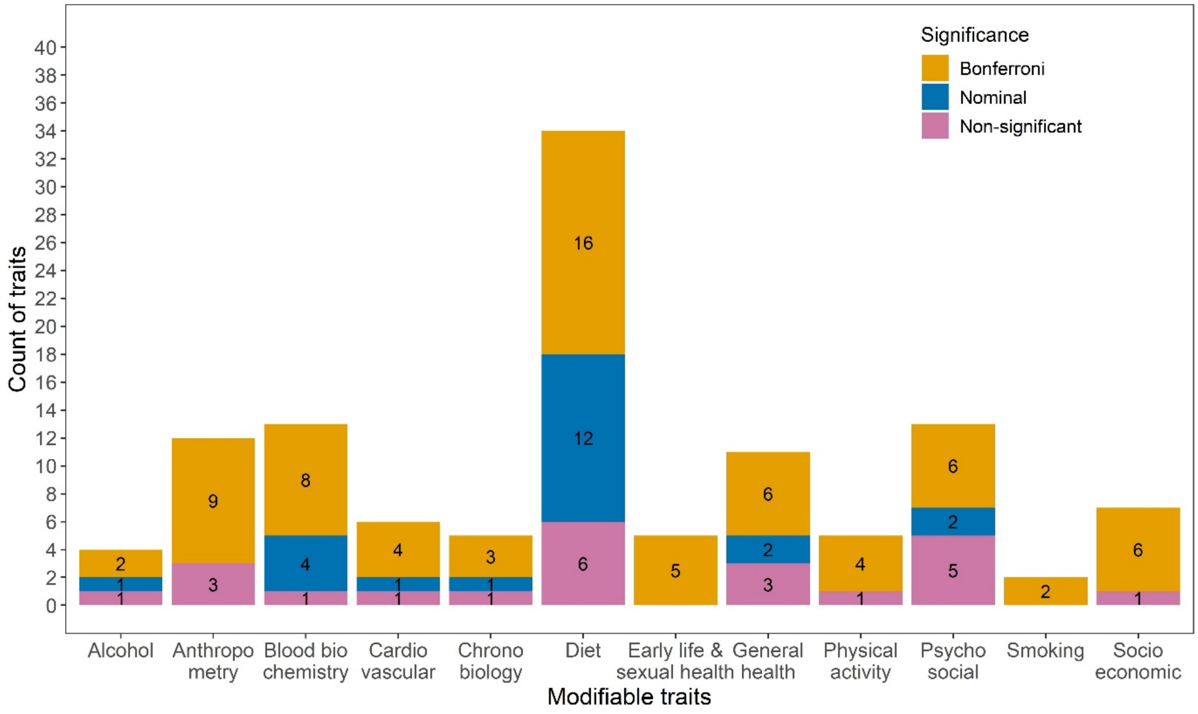


Bonferroni significance: $<4{\cdot}27\text{x}10^{-4}$; Nominal significance: $P<0{\cdot}05$; Non-significant: $P\geq0{\cdot}05$.

**Supplementary Figure 3. Association of leucocyte telomere length (LTL) with risk of 22 diseases across different scores for the primary healthy behaviour index.**

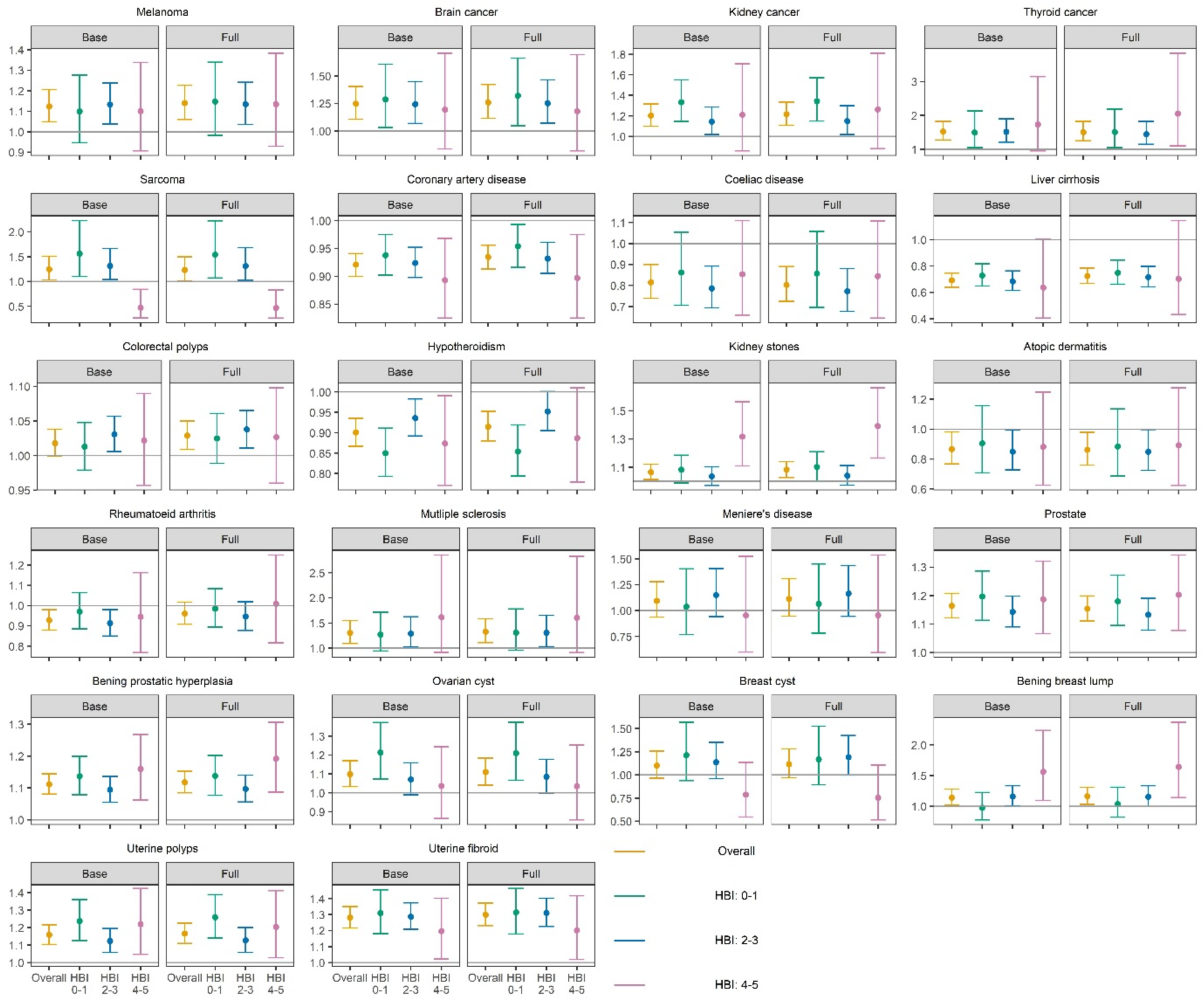


Each plot first shows the overall association of LTL disease with risk of disease and then the associations across three scores for the primary HBI: 0–1, 2–3, and 4–5 (all as hazard ratios per SD of longer LTL). The Base model is the model adjusted for age, sex, ethnicity and white blood cell count and the Full model is the model adjusted additionally for self–reported doctor diagnosed history of diabetes/ cancer/ hypertension/ vascular disease, highest qualification, insomnia, fed–up feelings, low–density cholesterol, C–reactive protein, and estimated glomerular filtration rate (CKD–EPI).

**Supplementary Figure 4. Years of life gained according to healthy behaviour index (HBI) groups with/without adjustment for leucocyte telomere length (LTL) as a mediator.**

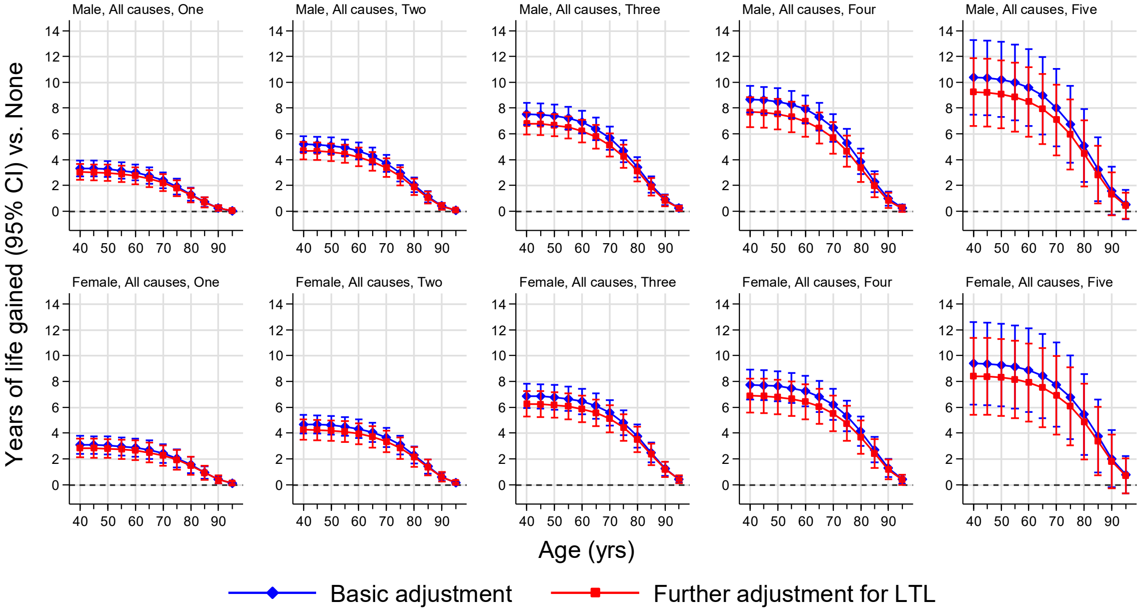


Years of life gained were estimated by applying hazard ratios (HRs) for cause–specific mortality calculated from UK Biobank data (specific to age–at–risk and stratified by sex and ethnic group) to population mortality rates for United Kingdom (UK) during year 2015 (by sex and 5–year age groups). Basic adjustment corresponded to age–at–risk specific HRs estimated in Cox regression (stratified by sex and ethnic group) and adjusted for white blood cell count. Further adjustment involved additional adjustment for LTL. The error bars are 95% confidence intervals (CI) and reflect uncertainty due to sampling variation in the HRs applied. The UK Biobank data included 329,826 participants and 19,140 deaths (comprising 4,001 vascular deaths, 10,301 cancer deaths, 4,710 non–vascular non–cancer deaths, and 128 deaths of unknown causes).

**Supplementary Figure 5. Mediation analysis to examine the proportion of the association of the healthy behaviour indices with CAD that may be mediated through leucocyte telomere length (LTL).**

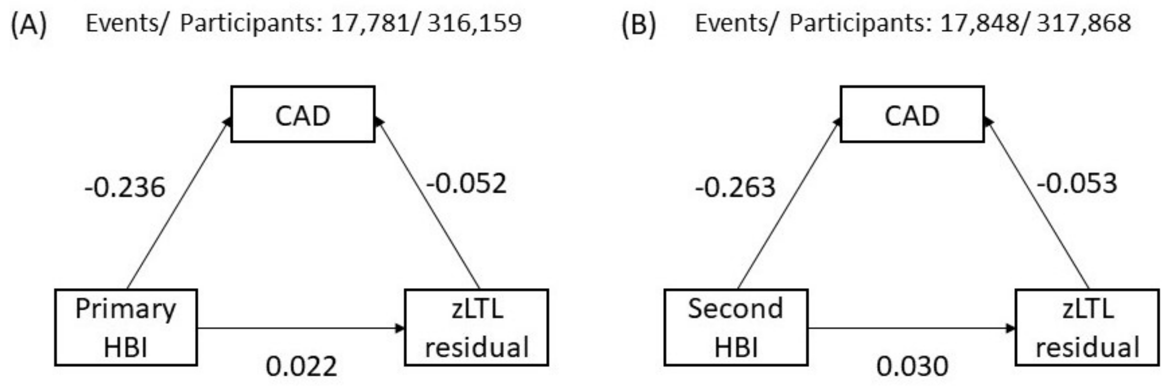


Structural equation models to investigate the proportion of the effect of healthy behaviours on coronary artery disease (CAD) that is mediated through telomere length. Model (A) is for the primary healthy behaviours index (HBI), model (B) is for the second index. zLTL residual is the standardised telomere length after removing the effect of age, sex, ethnicity and white blood cell count.

**Supplementary Table 1: Univariate model results for the association of modifiable traits with leucocyte telomere length (LTL).**

| Group | UKB field | Trait | | N | Mean(SD) / N(%) | Available data Beta (95% CI) | Pvalue | Equivalent years of age-related change in LTL | Imputed data (N=422,797 ) MI Beta (95% CI) | Pvalue | Equivalent years of age-related change in LTL |
|---|---|---|---|---|---|---|---|---|---|---|---|
| Alcohol | 1558 | Alcohol intake, frequency | | 422,407 | | *Global P:* | 0·486 | | *Global P:* | 0·475 | |
| | 1558 | | Never | | 86,777 (20·5%) | Reference | | | Reference | | |
| | 1558 | | Occassionally | | 98,010 (23·2%) | -0·002 (-0·016, 0·011) | 0·720 | -0·09 | -0·003 (-0·016, 0·011) | 0·706 | -0·11 |
| | 1558 | | 1-3 times/month | | 108,813 (25·8%) | 0·000 (-0·014, 0·013) | 0·954 | 0.00 | 0·000 (-0·014, 0·013) | 0·945 | 0.00 |
| | 1558 | | 1-2 times/week | | 46,768 (11·1%) | 0·005 (-0·007, 0·017) | 0·416 | 0·22 | 0·005 (-0·007, 0·017) | 0·417 | 0·22 |
| | 1558 | | 3-4 times/week | | 48,246 (11·4%) | 0·007 (-0·006, 0·019) | 0·292 | 0·30 | 0·007 (-0·006, 0·019) | 0·293 | 0·29 |
| | 1558 | | Daily | | 33,793 (8·0%) | 0·001 (-0·012, 0·014) | 0·863 | 0·04 | 0·001 (-0·012, 0·014) | 0·869 | 0·05 |
| | 1628 | Alcohol comparison, 10 years | | 385,725 | | *Global P:* | 9·78x10-18 | | *Global P:* | 1·46x10-16 | |
| | 1628 | | About the same | | 146,319 (37·9%) | Reference | | | Reference | | |
| | 1628 | | Less nowadays | | 175,332 (45·5%) | -0·031 (-0·037, -0·024) | 9·64x10-19 | -1·33 | -0·031 (-0·038, -0·024) | 2·77x10-18 | -1·34 |
| | 1628 | | More nowadays | | 64,074 (16·6%) | -0·018 (-0·027, -0·009) | 7·08x10-05 | -0·80 | -0·020 (-0·029, -0·011) | 1·00x10-05 | -0·89 |
| | 20117 | Drinking status | | 422,286 | | *Global P:* | 0·035 | | *Global P:* | 0·033 | |
| | 20117 | | Never | | 18,543 (4·4%) | Reference | | | Reference | | |
| | 20117 | | Previous | | 15,129 (3·6%) | -0·027 (-0·049, -0·006) | 0·011 | -1·17 | -0·028 (-0·049, -0·006) | 0·011 | -1·21 |
| | 20117 | | Current | | 388614 (92·0%) | -0·010 (-0·025, 0·005) | 0·205 | -0·43 | -0·010 (-0·025, 0·005) | 0·173 | -0·45 |
| | *Derived* | Alcohol intake, g/day | | 327,509 | 18·35 (16·92) | -0·001 (-0·002, -0·001) | 3·98x10-44 | -0·06 | -0·001 (-0·001, -0·001) | 1·65x10-37 | -0·05 |
| Anthropometry | 46 | Hand grip strength (left) | | 420,875 | 29·63 (11·25) | 0·018 (0·014, 0·022) | 9·75x10-16 | 0·78 | 0·018 (0·014, 0·023) | 5·16x10-16 | 0·79 |
| | 47 | Hand grip strength (right) | | 420,924 | 31·78 (11·19) | 0·017 (0·013, 0·021) | 3·22x10-14 | 0·74 | 0·017 (0·013, 0·022) | 1·71x10-14 | 0·75 |
| | 48 | Waist circumference | | 421,919 | 90·32 (13·32) | -0·020 (-0·023, -0·016) | 1·03x10-28 | -0·87 | -0·019 (-0·023, -0·016) | 1·86x10-28 | -0·84 |
| | 49 | Hip circumference | | 421,877 | 103·35 (9·01) | -0·012 (-0·015, -0·009) | 4·07x10-15 | -0·52 | -0·012 (-0·015, -0·009) | 7·83x10-15 | -0·51 |
| | 21002 | Weight | | 421,437 | 78·07 (15·69) | -0·014 (-0·018, -0·011) | 2·19x10-16 | -0·61 | -0·014 (-0·017, -0·011) | 4·42x10-16 | -0·61 |
| | 23099 | Body fat percentage | | 415,106 | 31·38 (8·48) | -0·035 (-0·039, -0·031) | 2·13x10-61 | -1·52 | -0·034 (-0·038, -0·029) | 2·09x10-52 | -1·46 |
| | 23100 | Whole body fat mass | | 414,635 | 24·78 (9·36) | -0·022 (-0·025, -0·019) | 2·39x10-42 | -0·96 | -0·021 (-0·024, -0·018) | 1·30x10-39 | -0·92 |
| | 23101 | Whole body fat-free mass | | 415,295 | 53·31 (11·44) | 0·002 (-0·004, 0·008) | 0·471 | 0·09 | 0·002 (-0·004, 0·008) | 0·471 | 0·09 |
| | 23102 | Whole body water mass | | 415,332 | 39·01 (8·37) | 0·002 (-0·004, 0·007) | 0·527 | 0·09 | 0·002 (-0·003, 0·008) | 0·432 | 0·10 |
| | 23104 | Body mass index | | 421,165 | 27·41 (4·70) | -0·023 (-0·026, -0·020) | 2·01x10-48 | -1.00 | -0·023 (-0·026, -0·019) | 6·87x10-48 | -0·98 |
| | 23105 | Basal metabolic rate | | 415,321 | 6,624 (1,353) | -0·004 (-0·009, 0·001) | 0·090 | -0·17 | -0·004 (-0·009, 0·001) | 0·139 | -0·16 |
| | *Derived* | Waist-hip circumference | | 421,837 | 0·87 (0·09) | -0·022 (-0·026, -0·018) | 1·52x10-25 | -0·96 | -0·021 (-0·025, -0·017) | 5·17x10-25 | -0·93 |
| | 30630 | Apolipoprotein A | | 368,909 | 1·54 (0·27) | -0·009 (-0·013, -0·006) | 8·93x10-08 | -0·39 | -0·009 (-0·012, -0·005) | 2·44x10-07 | -0·38 |
| | 30640 | Apolipoprotein B | | 403,228 | 1·03 (0·24) | 0·022 (0·019, 0·025) | 3·34x10-46 | 0·96 | 0·021 (0·018, 0·024) | 1·20x10-42 | 0·92 |
| | 30670 | Urea | | 405,029 | 5·39 (1·31) | 0·005 (0·001, 0·008) | 0·005 | 0·22 | 0·004 (0·001, 0·008) | 0·006 | 0·19 |
| | 30690 | Total cholesterol | | 405,304 | 5·69 (1·13) | 0·023 (0·019, 0·026) | 2·23x10-47 | 1.00 | 0·022 (0·019, 0·025) | 3·11x10-43 | 0·95 |

| | | | | | | | | | | | |
|---|---|---|---|---|---|---|---|---|---|---|---|
| Blood biochemistry | 30700 | eGFR | | 405,094 | 77·39 (75·02) | -0·040 (-0·052, -0·029) | 2·21x10-12 | -1·74 | -0·039 (-0·051, -0·028) | 2·73x10-11 | -1·71 |
| | 30710 | CRP | | 404,439 | 2·52 (3·70) | -0·023 (-0·026, -0·020) | 5·21x10-46 | -1.00 | -0·023 (-0·026, -0·020) | 1·74x10-44 | -0·99 |
| | 30740 | Glucose | | 370,704 | 5·11 (1·08) | -0·004 (-0·008, -0·001) | 0·006 | -0·17 | -0·005 (-0·008, -0·001) | 0·004 | -0·20 |
| | 30750 | HbA1c | | 402,742 | 36·07 (6·14) | -0·009 (-0·012, -0·006) | 2·24x10-08 | -0·39 | -0·009 (-0·012, -0·006) | 2·41x10-08 | -0·39 |
| | 30760 | HDL | | 370,963 | 1·45 (0·38) | -0·005 (-0·009, -0·002) | 0·004 | -0·22 | -0·004 (-0·007, 0·000) | 0·028 | -0·16 |
| | 30780 | LDL | | 404,553 | 3·56 (0·86) | 0·024 (0·021, 0·027) | 1·01x10-56 | 1·04 | 0·024 (0·021, 0·027) | 5·41x10-51 | 1·03 |
| | 30790 | Lipoprotein A | | 324,102 | 44·58 (49·13) | 0·005 (0·002, 0·008) | 0·003 | 0·22 | 0·004 (0·000, 0·007) | 0·026 | 0·17 |
| | 30870 | Triglycerides | | 404,972 | 1·74 (0·99) | 0·011 (0·008, 0·015) | 1·53x10-12 | 0·48 | 0·011 (0·007, 0·014) | 4·28x10-11 | 0·46 |
| | 30890 | Vitamin D | | 386,944 | 48·52 (20·81) | 0·001 (-0·002, 0·005) | 0·375 | 0·04 | 0·001 (-0·002, 0·004) | 0·508 | 0·05 |
| Cardiovascular | 102 | Pulse rate | | 399,424 | 69·33 (11·11) | -0·009 (-0·012, -0·006) | 1·44x10-08 | -0·39 | -0·008 (-0·012, -0·005) | 1·19x10-07 | -0·37 |
| | 4079 | Diastolic blood pressure | | 399,424 | 82·23 (10·03) | 0·007 (0·004, 0·010) | 6·19x10-06 | 0·30 | 0·007 (0·004, 0·010) | 3·66x10-06 | 0·31 |
| | 4080 | Systolic blood pressure | | 399,421 | 137·81 (18·42) | 0·009 (0·006, 0·013) | 1·46x10-08 | 0·39 | 0·009 (0·005, 0·012) | 3·09x10-07 | 0·38 |
| | 6032 | Maximum workload (fitness) | | 66,110 | 72·03 (35·61) | 0·013 (0·004, 0·022) | 0·004 | 0·57 | 0·020 (0·016, 0·023) | 1·61x10-19 | 0·85 |
| | 6033 | Maximum heart rate (fitness) | | 66,094 | 110·20 (19·60) | 0·001 (-0·007, 0·008) | 0·880 | 0·04 | 0·003 (-0·002, 0·007) | 0·231 | 0·11 |
| | *Derived* | Pulse pressure | | 399,421 | 55·57 (13·45) | 0·008 (0·004, 0·011) | 1·03x10-05 | 0·35 | 0·006 (0·003, 0·010) | 4·13x10-04 | 0·27 |
| Chronobiology | 1160 | Sleep duration | | 420,164 | 7·15 (1·07) | 0·000 (-0·003, 0·003) | 0·876 | 0.00 | 0·000 (-0·003, 0·003) | 0·876 | 0.00 |
| | 1170 | Getting up in morning | | 422,066 | | *Global P:* | 1·54x10-04 | | *Global P:* | 1·50x10-04 | |
| | 1170 | | Not at all easy | | 16,585 (3·9%) | Reference | | | Reference | | |
| | 1170 | | Not very easy | | 59,231 (14·0%) | 0·018 (0·001, 0·034) | 0·038 | 0·78 | 0·018 (0·001, 0·035) | 0·037 | 0·78 |
| | 1170 | | Fairly easy | | 209,653 (49·7%) | 0·020 (0·005, 0·036) | 0·009 | 0·87 | 0·021 (0·005, 0·036) | 0·009 | 0·89 |
| | 1170 | | Very easy | | 136,597 (32·4%) | 0·007 (-0·009, 0·023) | 0·394 | 0·30 | 0·007 (-0·009, 0·023) | 0·392 | 0·30 |
| | 1190 | Day nap | | 422,070 | | *Global P:* | 1·95x10-09 | | *Global P:* | 2·06x10-09 | |
| | 1190 | | Never/rarely | | 237,109 (56·2%) | Reference | | | Reference | | |
| | 1190 | | Sometimes | | 162,339 (38·5%) | -0·016 (-0·023, -0·010) | 2·76x10-07 | -0·70 | -0·016 (-0·023, -0·010) | 2·89x10-07 | -0·71 |
| | 1190 | | Usually | | 22,622 (5·3%) | -0·032 (-0·046, -0·019) | 2·65x10-06 | -1·39 | -0·032 (-0·046, -0·019) | 2·66x10-06 | -1·40 |
| | 1200 | Insomnia | | 422,390 | | *Global P:* | 1·00x10-09 | | *Global P:* | 1·10x10-09 | |
| | 1200 | | Never/rarely | | 102,340 (24·2%) | Reference | | | Reference | | |
| | 1200 | | Sometimes | | 201,451 (47·7%) | -0·011 (-0·018, -0·003) | 0·005 | -0·48 | -0·011 (-0·018, -0·003) | 0·005 | -0·46 |
| | 1200 | | Usually | | 118,599 (28·1%) | -0·027 (-0·035, -0·018) | 2·78x10-10 | -1·17 | -0·027 (-0·035, -0·018) | 3·06x10-10 | -1·15 |
| | 1220 | Narcolepsy | | 420,565 | | *Global P:* | 4·89x10-04 | | *Global P:* | 7·56x10-04 | |
| | 1220 | | Never/rarely | | 319,550 (76·0%) | Reference | | | Reference | | |
| | 1220 | | Sometimes | | 89,228 (21·2%) | -0·007 (-0·014, 0·001) | 0·073 | -0·30 | -0·006 (-0·014, 0·001) | 0·082 | -0·28 |
| | 1220 | | Often | | 11,752 (2·8%) | -0·036 (-0·054, -0·018) | 8·11x10-05 | -1·57 | -0·036 (-0·054, -0·018) | 7·46x10-05 | -1·57 |
| | 1220 | | All of the time | | 35 (0·0%) | 0·076 (-0·247, 0·398) | 0·645 | 3·30 | 0·087 (-0·244, 0·417) | 0·606 | 3·77 |
| | 1289 | Cooked vegetable intake | | 408,950 | 2·74 (1·59) | 0·006 (0·003, 0·009) | 5·43x10-05 | 0·26 | 0·006 (0·003, 0·009) | 3·67x10-05 | 0·28 |
| | 1299 | Raw vegetable intake | | 397,383 | 2·24 (1·88) | 0·008 (0·005, 0·011) | 3·21x10-07 | 0·35 | 0·008 (0·005, 0·011) | 1·41x10-07 | 0·35 |
| | 1309 | Fresh fruit intake | | 407,512 | 2·27 (1·45) | 0·016 (0·013, 0·019) | 3·69x10-26 | 0·70 | 0·016 (0·013, 0·019) | 2·95x10-26 | 0·71 |
| | 1319 | Dried fruit intake | | 384,574 | 0·86 (1·54) | 0·025 (0·022, 0·028) | 9·48x10-54 | 1·09 | 0·024 (0·021, 0·027) | 1·36x10-48 | 1·03 |

| | | | | | | | | | | |
|---|---|---|---|---|---|---|---|---|---|---|
| 1329 | Oily fish intake | | 420,294 | | *Global P:* | 1·56x10-28 | | *Global P:* | 1·93x10-28 | |
| 1329 | | Never | | 45,794 (10·9%) | Reference | | | Reference | | |
| 1329 | | Less than once a week | | 139,316 (33·1%) | 0·040 (0·029, 0·050) | 5·51x10-14 | 1·74 | 0·040 (0·029, 0·050) | 3·92x10-14 | 1·73 |
| 1329 | | Once a week | | 159,100 (37·9%) | 0·051 (0·041, 0·061) | 1·70x10-22 | 2·22 | 0·051 (0·041, 0·061) | 8·60x10-23 | 2·22 |
| 1329 | | 2-4 times a week | | 72,069 (17·1%) | 0·064 (0·052, 0·075) | 3·51x10-27 | 2·78 | 0·064 (0·052, 0·075) | 1·97x10-27 | 2·77 |
| 1329 | | 5+ times a week | | 4,015 (1·0%) | 0·085 (0·054, 0·116) | 1·19x10-07 | 3·70 | 0·083 (0·051, 0·114) | 3·07x10-07 | 3·59 |
| 1339 | Non-oily fish intake | | 420,586 | | *Global P:* | 0·025 | | *Global P:* | 0·026 | |
| 1339 | | Never | | 19,725 (4·7%) | Reference | | | Reference | | |
| 1339 | | Less than once a week | | 122,050 (29·0%) | 0·020 (0·005, 0·035) | 0·007 | 0·87 | 0·021 (0·006, 0·036) | 0·006 | 0·91 |
| 1339 | | Once a week | | 209,634 (49·8%) | 0·019 (0·004, 0·033) | 0·011 | 0·83 | 0·020 (0·005, 0·034) | 0·008 | 0·85 |
| 1339 | | 2-4 times a week | | 66,521 (15·8%) | 0·019 (0·003, 0·034) | 0·018 | 0·83 | 0·020 (0·004, 0·036) | 0·013 | 0·87 |
| 1339 | | 5+ times a week | | 2,656 (0·7%) | 0·055 (0·016, 0·095) | 0·006 | 2·39 | 0·052 (0·012, 0·092) | 0·011 | 2·26 |
| 1349 | Processed meat intake | | 421,808 | | *Global P:* | 7·61x10-21 | | *Global P:* | 1·35x10-21 | |
| 1349 | | Never | | 39,127 (9·3%) | Reference | | | Reference | | |
| 1349 | | Less than once a week | | 128,413 (30·4%) | -0·022 (-0·033, -0·011) | 1·23x10-04 | -0·96 | -0·021 (-0·032, -0·010) | 1·54x10-04 | -0·93 |
| 1349 | | Once a week | | 123,038 (29·2%) | -0·042 (-0·053, -0·031) | 2·70x10-13 | -1·83 | -0·041 (-0·053, -0·030) | 4·45x10-13 | -1·80 |
| 1349 | | 2-4 times a week | | 114,385 (27·1%) | -0·050 (-0·062, -0·039) | 1·16x10-17 | -2·17 | -0·050 (-0·061, -0·038) | 2·20x10-17 | -2·16 |
| 1349 | | 5+ times a week | | 16,845 (4·0%) | -0·054 (-0·072, -0·036) | 2·62x10-09 | -2·35 | -0·054 (-0·072, -0·036) | 2·81x10-09 | -2·36 |
| 1359 | Poultry intake | | 421,939 | | *Global P:* | 1·18x10-07 | | *Global P:* | 1·60x10-07 | |
| 1359 | | Never | | 21,494 (5·1%) | Reference | | | Reference | | |
| 1359 | | Less than once a week | | 45,438 (10·8%) | -0·028 (-0·044, -0·012) | 6·35x10-04 | -1·22 | -0·027 (-0·043, -0·011) | 7·49x10-04 | -1·19 |
| 1359 | | Once a week | | 151,450 (35·9%) | -0·034 (-0·048, -0·020) | 1·55x10-06 | -1·48 | -0·034 (-0·048, -0·020) | 1·94x10-06 | -1·48 |
| 1359 | | 2-4 times a week | | 193,971 (46·0%) | -0·041 (-0·055, -0·027) | 6·29x10-09 | -1·78 | -0·040 (-0·054, -0·027) | 8·93x10-09 | -1·76 |
| 1359 | | 5+ times a week | | 9,586 (2·2%) | -0·044 (-0·067, -0·020) | 2·89x10-04 | -1·91 | -0·043 (-0·067, -0·020) | 2·93x10-04 | -1·89 |
| 1369 | Beef intake | | 420,909 | | *Global P:* | 5·60x10-08 | | *Global P:* | 5·70x10-08 | |
| 1369 | | Never | | 46,538 (11·1%) | Reference | | | Reference | | |
| 1369 | | Less than once a week | | 191,645 (45·5%) | -0·020 (-0·031, -0·010) | 7·65x10-05 | -0·87 | -0·020 (-0·030, -0·010) | 1·12x10-04 | -0·87 |
| 1369 | | Once a week | | 133,926 (31·8%) | -0·031 (-0·041, -0·020) | 1·15x10-08 | -1·35 | -0·031 (-0·041, -0·020) | 1·31x10-08 | -1·34 |
| 1369 | | 2-4 times a week | | 47,710 (11·3%) | -0·033 (-0·046, -0·020) | 4·21x10-07 | -1·43 | -0·033 (-0·045, -0·020) | 4·53x10-07 | -1·42 |
| 1369 | | 5+ times a week | | 1,090 (0·3%) | -0·051 (-0·110, 0·007) | 0·087 | -2·22 | -0·054 (-0·112, 0·003) | 0·064 | -2·37 |
| 1379 | Lamb intake | | 419,924 | | *Global P:* | 0·761 | | *Global P:* | 0·708 | |
| 1379 | | Never | | 74,379 (17·7%) | Reference | | | Reference | | |
| 1379 | | Less than once a week | | 237,891 (56·7%) | 0·002 (-0·006, 0·010) | 0·621 | 0·09 | 0·002 (-0·006, 0·010) | 0·615 | 0·09 |
| 1379 | | Once a week | | 94,754 (22·5%) | -0·003 (-0·012, 0·007) | 0·594 | -0·13 | -0·003 (-0·012, 0·007) | 0·593 | -0·11 |
| 1379 | | 2-4 times a week | | 12,442 (3·0%) | 0·001 (-0·018, 0·019) | 0·953 | 0·04 | 0·001 (-0·018, 0·019) | 0·955 | 0·02 |
| 1379 | | 5+ times a week | | 458 (0·1%) | -0·026 (-0·116, 0·064) | 0·573 | -1·13 | -0·033 (-0·118, 0·053) | 0·452 | -1·43 |
| 1389 | Pork intake | | 420,120 | | *Global P:* | 0·006 | | *Global P:* | 0·008 | |
| 1389 | | Never | | 72,258 (17·2%) | Reference | | | | | |

| | | | | | | | | | | | |
|---|---|---|---|---|---|---|---|---|---|---|---|
| | 1389 | | Less than once a week | | 238,916 (56·9%) | -0·007 (-0·015, 0·001) | 0·100 | -0·30 | -0·006 (-0·015, 0·002) | 0·125 | -0·28 |
| | 1389 | | Once a week | | 93,869 (22·3%) | -0·017 (-0·027, -0·008) | 4·00x10-04 | -0·74 | -0·017 (-0·027, -0·007) | 5·30x10-04 | -0·74 |
| | 1389 | | 2-4 times a week | | 14,466 (3·4%) | -0·011 (-0·028, 0·007) | 0·232 | -0·48 | -0·010 (-0·028, 0·007) | 0·250 | -0·45 |
| | 1389 | | 5+ times a week | | 611 (0·2%) | -0·046 (-0·124, 0·031) | 0·243 | -2.00 | -0·042 (-0·118, 0·033) | 0·272 | -1·84 |
| | 1408 | Cheese intake | | 411,632 | | *Global P:* | 1·08x10-24 | | *Global P:* | 9·56x10-25 | |
| | 1408 | | Never | | 11,265 (2·7%) | Reference | | | Reference | | |
| | 1408 | | Less than once a week | | 71,024 (17·3%) | 0·013 (-0·007, 0·032) | 0·193 | 0·57 | 0·013 (-0·006, 0·032) | 0·182 | 0·57 |
| | 1408 | | Once a week | | 87,902 (21·3%) | 0·011 (-0·008, 0·030) | 0·243 | 0·48 | 0·012 (-0·007, 0·031) | 0·227 | 0·51 |
| | 1408 | | 2-4 times a week | | 186,658 (45·4%) | 0·021 (0·002, 0·039) | 0·027 | 0·91 | 0·021 (0·003, 0·040) | 0·023 | 0·93 |
| | 1408 | | 5+ times a week | | 54,783 (13·3%) | 0·063 (0·043, 0·083) | 4·55x10-10 | 2·74 | 0·063 (0·044, 0·083) | 3·04x10-10 | 2·76 |
| Diet | 1418 | Milk type | | 422,450 | | *Global P:* | 3·45x10-06 | | *Global P:* | 3·33x10-06 | |
| | 1418 | | Never/rarely | | 14,115 (3·3%) | Reference | | | Reference | | |
| | 1418 | | Full cream | | 29,193 (6·9%) | -0·011 (-0·031, 0·008) | 0·266 | -0·48 | -0·011 (-0·031, 0·008) | 0·255 | -0·49 |
| | 1418 | | Semi-skimmed | | 272,463 (64·5%) | -0·003 (-0·019, 0·014) | 0·742 | -0·12 | -0·003 (-0·020, 0·013) | 0·712 | -0·13 |
| | 1418 | | Skimmed | | 84,692 (20·1%) | -0·002 (-0·020, 0·015) | 0·803 | -0·10 | -0·003 (-0·020, 0·015) | 0·770 | -0·11 |
| | 1418 | | Soya | | 16,551 (3·9%) | 0·040 (0·018, 0·062) | 3·40x10-04 | 1·74 | 0·040 (0·018, 0·062) | 3·71x10-04 | 1·73 |
| | 1418 | | Other type of milk | | 5,436 (1·3%) | -0·005 (-0·036, 0·025) | 0·741 | -0·22 | -0·005 (-0·036, 0·025) | 0·724 | -0·24 |
| | 1428 | Spread type | | 421,983 | | *Global P:* | 0·007 | | *Global P:* | 0·007 | |
| | 1428 | | Rarely use spread | | 45,785 (10·9%) | Reference | | | Reference | | |
| | 1428 | | Butter | | 153,640 (36·4%) | -0·013 (-0·023, -0·003) | 0·014 | -0·57 | -0·013 (-0·023, -0·003) | 0·014 | -0·55 |
| | 1428 | | Flora/Benecol | | 2,392 (0·6%) | -0·006 (-0·046, 0·034) | 0·773 | -0·26 | -0·005 (-0·045, 0·035) | 0·801 | -0·22 |
| | 1428 | | Other spread | | 220,166 (52·1%) | -0·017 (-0·027, -0·007) | 5·80x10-04 | -0·74 | -0·017 (-0·027, -0·007) | 5·76x10-04 | -0·75 |
| | 1448 | Bread type | | 407,749 | | *Global P:* | 1·25x10-53 | | *Global P:* | 1·36x10-52 | |
| | 1448 | | White | | 107,314 (26·3%) | Reference | | | Reference | | |
| | 1448 | | Brown | | 51,406 (12·6%) | 0·032 (0·021, 0·042) | 1·39x10-09 | 1·39 | 0·031 (0·021, 0·042) | 1·82x10-09 | 1·37 |
| | 1448 | | Wholemeal | | 231,557 (56·8%) | 0·057 (0·050, 0·065) | 1·90x10-55 | 2·48 | 0·057 (0·050, 0·064) | 2·65x10-55 | 2·48 |
| | 1448 | | Other type | | 17,472 (4·3%) | 0·044 (0·028, 0·060) | 3·78x10-08 | 1·91 | 0·045 (0·029, 0·061) | 3·03x10-08 | 1·95 |
| | 1468 | Cereal type | | 347,253 | | *Global P:* | 2·92x10-54 | | *Global P:* | 7·38x10-41 | |
| | 1468 | | Other | | 67,901 (19·6%) | Reference | | | Reference | | |
| | 1468 | | Bran cereal | | 58,374 (16·8%) | 0·026 (0·016, 0·037) | 1·64x10-06 | 1·15 | 0·027 (0·017, 0·038) | 7·01x10-07 | 1·19 |
| | 1468 | | Biscuit cereal | | 61,247 (17·6%) | 0·004 (-0·007, 0·014) | 0·486 | 0·16 | 0·006 (-0·004, 0·016) | 0·254 | 0·26 |
| | 1468 | | Oat cereal | | 88,946 (25·6%) | 0·034 (0·024, 0·044) | 1·20x10-11 | 1·48 | 0·035 (0·024, 0·045) | 7·98x10-10 | 1·51 |
| | 1468 | | Muesli | | 70,785 (20·4%) | 0·075 (0·064, 0·085) | 1·11x10-45 | 3·24 | 0·070 (0·060, 0·080) | 3·35x10-29 | 3·05 |
| | 1478 | Added salt | | 422,709 | | *Global P:* | 1·74x10-35 | | *Global P:* | 2·12x10-35 | |
| | 1478 | | Never/rarely | | 234,081 (55·4%) | Reference | | | Reference | | |
| | 1478 | | Sometimes | | 118,724 (28·1%) | -0·024 (-0·031, -0·018) | 2·43x10-12 | -1·04 | -0·024 (-0·031, -0·018) | 2·55x10-12 | -1·06 |
| | 1478 | | Usually | | 49,402 (11·7%) | -0·038 (-0·047, -0·028) | 4·44x10-15 | -1·65 | -0·038 (-0·047, -0·028) | 4·77x10-15 | -1·64 |
| | 1478 | | Always | | 20,502 (4·9%) | -0·073 (-0·087, -0·059) | 1·15x10-24 | -3·17 | -0·073 (-0·087, -0·059) | 1·32x10-24 | -3·17 |

| | | | | | | | | | | |
|---|---|---|---|---|---|---|---|---|---|---|
| | 1488 | Tea intake | 408,734 | 3·48 (2·68) | -0·003 (-0·006, 0·000) | 0·026 | -0·13 | -0·004 (-0·007, -0·001) | 0·017 | -0·16 |
| | 1498 | Coffee intake | 391,072 | 2·12 (2·00) | -0·003 (-0·006, 0·000) | 0·089 | -0·13 | -0·002 (-0·005, 0·001) | 0·221 | -0·08 |
| | 1528 | Water intake | 391,350 | 2·87 (2·13) | 0·009 (0·006, 0·012) | 2·71x10-08 | 0·39 | 0·008 (0·005, 0·012) | 1·27x10-07 | 0·37 |
| | 2654 | Vegetable spread type | 219,677 | | *Global P:* | 7·44x10-18 | | *Global P:* | 4·12x10-15 | |
| | 2654 | Olive oil based spread | | 54,213 (24·7%) | Reference | | | Reference | | |
| | 2654 | Flora/Benecol | | 33,876 (15·4%) | -0·008 (-0·021, 0·005) | 0·238 | -0·35 | -0·002 (-0·014, 0·011) | 0·789 | -0·08 |
| | 2654 | Soft (tub) margarine | | 28,194 (12·8%) | -0·066 (-0·080, -0·052) | 3·32x10-20 | -2·88 | -0·063 (-0·077, -0·049) | 2·99x10-18 | -2·73 |
| | 2654 | Hard (block) margarine | | 341 (0·2%) | 0·011 (-0·093, 0·114) | 0·838 | 0·47 | 0·008 (-0·033, 0·049) | 0·697 | 0·35 |
| | 2654 | Polyunsaturated/sunflower | | 73,234 (33·3%) | -0·022 (-0·033, -0·012) | 4·87x10-05 | -0·97 | -0·022 (-0·033, -0·011) | 5·99x10-05 | -0·96 |
| | 2654 | Other low/reduced fat spread | | 21,789 (9·9%) | -0·028 (-0·044, -0·013) | 2·62x10-04 | -1·24 | -0·018 (-0·028, -0·009) | 1·86x10-04 | -0·79 |
| | 2654 | Other type of spread | | 8,030 (3·7%) | -0·024 (-0·047, -0·001) | 0·037 | -1·05 | -0·015 (-0·035, 0·006) | 0·169 | -0·63 |
| | 6155 | Vitamin supplement | 434,860 | | | | | | | |
| | 6155_1 | Vitamin A (yes vs no) | | 8,799 (2·0%) | 0·015 (-0·006, 0·036) | 0·159 | 0·65 | 0·015 (-0·006, 0·036) | 0·159 | 0·65 |
| | 6155_2 | Vitamin B (yes vs no) | | 18,809 (4·3%) | -0·004 (-0·018, 0·011) | 0·599 | -0·17 | -0·004 (-0·018, 0·011) | 0·610 | -0·16 |
| | 6155_3 | Vitamin C (yes vs no) | | 38,414 (8·8%) | 0·016 (0·006, 0·027) | 0·002 | 0·70 | 0·016 (0·006, 0·027) | 0·002 | 0·71 |
| | 6155_4 | Vitamin D (yes vs no) | | 17,567 (4·0%) | 0·021 (0·006, 0·036) | 0·005 | 0·91 | 0·021 (0·006, 0·036) | 0·006 | 0·92 |
| | 6155_5 | Vitamin E (yes vs no) | | 13,338 (3·1%) | 0·019 (0·002, 0·036) | 0·030 | 0·83 | 0·019 (0·002, 0·036) | 0·029 | 0·82 |
| | 6155_6 | Folic acid (vit B9) (yes vs no) | | 9,880 (2·3%) | -0·022 (-0·042, -0·002) | 0·030 | -0·96 | -0·022 (-0·041, -0·002) | 0·033 | -0·94 |
| | 6155_7 | Multivitamins (yes vs no) | | 95,930 (22·1%) | 0·011 (0·004, 0·018) | 0·002 | 0·48 | 0·011 (0·004, 0·018) | 0·002 | 0·49 |
| | 6179 | Mineral supplement | 435,861 | | | | | | | |
| | 6179_1 | Fish oil (yes vs no) | | 136,848 (31·4%) | 0·006 (0·000, 0·013) | 0·057 | 0·26 | 0·006 (0·000, 0·013) | 0·054 | 0·28 |
| | 6179_2 | Glucosamine (yes vs no) | | 83,394 (19·1%) | 0·014 (0·007, 0·022) | 1·93x10-04 | 0·61 | 0·014 (0·007, 0·022) | 1·83x10-04 | 0·63 |
| | 6179_3 | Calcium (yes vs no) | | 30,660 (7·0%) | 0·012 (0·000, 0·024) | 0·048 | 0·52 | 0·012 (0·000, 0·024) | 0·045 | 0·52 |
| | 6179_4 | Zinc (yes vs no) | | 18,045 (4·1%) | 0·018 (0·004, 0·033) | 0·014 | 0·78 | 0·018 (0·004, 0·033) | 0·015 | 0·80 |
| | 6179_5 | Iron (yes vs no) | | 14,515 (3·3%) | 0·012 (-0·005, 0·028) | 0·157 | 0·52 | 0·012 (-0·005, 0·028) | 0·165 | 0·51 |
| | 6179_6 | Selenium (yes vs no) | | 10,516 (2·4%) | 0·028 (0·008, 0·047) | 0·005 | 1·22 | 0·028 (0·009, 0·047) | 0·004 | 1·21 |
| Early life and sexual health | 1677 | Breastfed as a baby | 323,147 | 233,570 (72·3%) | 0·034 (0·027, 0·042) | 2·39x10-18 | 1·48 | 0·029 (0·022, 0·037) | 7·91x10-12 | 1·28 |
| | 1787 | Maternal smoking around birth | 364,690 | 107,180 (29·4%) | -0·054 (-0·061, -0·047) | 2·94x10-52 | -2·35 | -0·050 (-0·057, -0·043) | 9·02x10-44 | -2·16 |
| | 2139 | Age first sexual intercourse | 367,951 | 19·10 (3·66) | 0·035 (0·032, 0·038) | 1·54x10-99 | 1·52 | 0·033 (0·030, 0·036) | 5·17x10-58 | 1·44 |
| | 2754 | Age first live birth | 153,938 | 25·37 (4·59) | 0·045 (0·040, 0·050) | 5·87x10-69 | 1·96 | 0·034 (0·029, 0·038) | 1·10x10-15 | 1·47 |
| | 2764 | Age last live birth | 153,638 | 30·31 (4·86) | 0·032 (0·027, 0·037) | 1·23x10-36 | 1·39 | 0·023 (0·019, 0·028) | 5·51x10-14 | 1·02 |
| alth | 78 | Bone mineral density | 237,905 | -0·34 (1·19) | 0·001 (-0·003, 0·005) | 0·605 | 0·04 | 0·000 (-0·003, 0·004) | 0·874 | 0.00 |
| | 2178 | Overall health status | 420,835 | | *Global P:* | 2·13x10-70 | | *Global P:* | 1·63x10-70 | |
| | 2178 | Excellent | | 69,612 (16·5%) | Reference | | | Reference | | |
| | 2178 | Good | | 244,178 (58·0%) | -0·030 (-0·039, -0·022) | 3·80x10-13 | -1·3 | -0·030 (-0·039, -0·022) | 3·93x10-13 | -1·32 |
| | 2178 | Fair | | 88,236 (21·0%) | -0·068 (-0·078, -0·058) | 2·86x10-42 | -2·96 | -0·068 (-0·078, -0·058) | 1·41x10-42 | -2·97 |
| | 2178 | Poor | | 18,809 (4·5%) | -0·120 (-0·136, -0·104) | 5·97x10-50 | -5·22 | -0·120 (-0·135, -0·104) | 7·65x10-50 | -5·2 |
| | 3064 | Peak expiratory flow | 385,708 | 408·89 (125·34) | 0·024 (0·020, 0·028) | 2·54x10-30 | 1·04 | 0·024 (0·021, 0·028) | 2·38x10-34 | 1·06 |

| | | | | | | | | | | | |
|---|---|---|---|---|---|---|---|---|---|---|---|
| General he | 6149 | Dental problems | | 421,450 | | | | | | | |
| | 6149_1 | | Mouth ulcers (yes vs no) | | 42,691 (10·1%) | 0·014 (0·005, 0·024) | 0·004 | 0·63 | 0·015 (0·005, 0·024) | 0·004 | 0·63 |
| | 6149_2 | | Painful gums (yes vs no) | | 12,855 (3·1%) | -0·023 (-0·041, -0·006) | 0·007 | -1·02 | -0·023 (-0·041, -0·006) | 0·007 | -1·02 |
| | 6149_3 | | Bleeding gums (yes vs no) | | 56,496 (13·4%) | 0·018 (0·009, 0·027) | 5·48x10-05 | 0·78 | 0·018 (0·009, 0·026) | 6·02x10-05 | 0·77 |
| | 6149_4 | | Loose teeth (yes vs no) | | 18,402 (4·4%) | -0·061 (-0·076, -0·047) | 1·02x10-16 | -2·66 | -0·061 (-0·075, -0·047) | 1·38x10-16 | -2·65 |
| | 6149_5 | | Toothache (yes vs no) | | 18,831 (4·5%) | 0·000 (-0·014, 0·015) | 0·953 | 0·02 | 0·000 (-0·014, 0·015) | 0·952 | 0·02 |
| | 6149_6 | | Dentures (yes vs no) | | 69,920 (16·6%) | -0·063 (-0·071, -0·054) | 1·94x10-50 | -2·72 | -0·062 (-0·071, -0·054) | 3·74x10-50 | -2·71 |
| | 30520 | Potassium in urine | | 409,917 | 63·10 (33·63) | 0·002 (-0·001, 0·005) | 0·118 | 0·09 | 0·002 (-0·001, 0·005) | 0·131 | 0·10 |
| | 30530 | Sodium in urine | | 409,928 | 77·40 (44·03) | -0·007 (-0·010, -0·004) | 3·68x10-06 | -0·30 | -0·007 (-0·010, -0·004) | 5·58x10-06 | -0·31 |
| Physical activity | 924 | Walking pace | | 419,819 | | *Global P:* | 8·95x10-73 | | *Global P:* | 3·00x10-74 | |
| | 924 | | Slow pace | | 33,821 (8·1%) | Reference | | | Reference | | |
| | 924 | | Average pace | | 221,482 (52·8%) | 0·068 (0·057, 0·080) | 5·72x10-33 | 2·96 | 0·069 (0·058, 0·080) | 3·21x10-34 | 3.00 |
| | 924 | | Brisk pace | | 164,516 (39·1%) | 0·103 (0·092, 0·115) | 6·23x10-68 | 4·48 | 0·104 (0·092, 0·115) | 4·97x10-70 | 4·51 |
| | 22037 | METs walking | | 342,455 | 1,038·14 (1,085·23) | -0·009 (-0·012, -0·005) | 3·02x10-07 | -0·39 | -0·006 (-0·009, -0·003) | 4·81x10-05 | -0·28 |
| | 22038 | METs moderate activity | | 342,455 | 931·92 (1,224·55) | -0·009 (-0·012, -0·006) | 1·27x10-07 | -0·39 | -0·006 (-0·009, -0·003) | 5·89x10-05 | -0·27 |
| | 22039 | METs vigorous activity | | 342,455 | 666·76 (1,140·98) | 0·011 (0·008, 0·015) | 8·58x10-12 | 0·48 | 0·014 (0·010, 0·017) | 2·29x10-16 | 0·59 |
| | 22040 | Total METs | | 342,455 | 2,636·47 (2,657·64) | 0·003 (-0·001, 0·006) | 0·114 | 0·13 | 0·006 (0·003, 0·009) | 4·01x10-05 | 0·28 |
| Psychosocial | 1920 | Mood swings | | 411,691 | 186,961 (45·4%) | -0·028 (-0·034, -0·022) | 9·72x10-20 | -1·22 | -0·027 (-0·033, -0·021) | 2·84x10-18 | -1·16 |
| | 1930 | Miserableness | | 415,000 | 176,981 (42·7%) | -0·021 (-0·027, -0·015) | 2·63x10-11 | -0·91 | -0·020 (-0·026, -0·014) | 6·28x10-11 | -0·89 |
| | 1940 | Irritability | | 403,170 | 113,003 (28·0%) | -0·009 (-0·016, -0·002) | 0·008 | -0·39 | -0·009 (-0·016, -0·003) | 0·006 | -0·40 |
| | 1950 | Sensitivity/ hurt feelings | | 410,054 | 227,262 (55·4%) | -0·015 (-0·021, -0·009) | 2·26x10-06 | -0·65 | -0·014 (-0·020, -0·008) | 4·56x10-06 | -0·62 |
| | 1960 | Fed-up feelings | | 413,210 | 167,429 (40·5%) | -0·033 (-0·039, -0·027) | 7·30x10-26 | -1·43 | -0·032 (-0·038, -0·026) | 7·26x10-25 | -1·39 |
| | 1970 | Nervous feelings | | 411,223 | 96,822 (23·5%) | 0·000 (-0·007, 0·007) | 0·899 | 0.00 | 0·000 (-0·007, 0·007) | 0·982 | 0.00 |
| | 1980 | Worrier/ anxious feelings | | 411,254 | 232,064 (56·4%) | 0·001 (-0·006, 0·007) | 0·867 | 0·04 | 0·001 (-0·006, 0·007) | 0·859 | 0·02 |
| | 1990 | Tense/ 'highly strung' | | 407,212 | 72,141 (17·7%) | -0·011 (-0·019, -0·003) | 0·008 | -0·48 | -0·010 (-0·018, -0·003) | 0·008 | -0·45 |
| | 2000 | Worry too long after embarrassment | | 404,972 | 192,982 (47·7%) | 0·000 (-0·006, 0·006) | 0·914 | 0.00 | 0·000 (-0·006, 0·006) | 0·992 | 0.00 |
| | 2020 | Loneliness, isolation | | 415,520 | 76,486 (18·4%) | -0·018 (-0·025, -0·010) | 5·77x10-06 | -0·78 | -0·017 (-0·025, -0·010) | 7·71x10-06 | -0·76 |
| | 2030 | Guilty feelings | | 410,889 | 118,149 (28·8%) | -0·006 (-0·012, 0·001) | 0·094 | -0·26 | -0·007 (-0·013, 0·000) | 0·054 | -0·28 |
| | 2040 | Risk taking | | 406,629 | 110,057 (27·1%) | -0·005 (-0·012, 0·002) | 0·162 | -0·22 | -0·005 (-0·012, 0·002) | 0·173 | -0·21 |
| | 20127 | Neuroticism score | | 339,183 | 4·11 (3·27) | -0·009 (-0·012, -0·006) | 1·29x10-07 | -0·39 | -0·009 (-0·012, -0·006) | 2·15x10-09 | -0·40 |
| Smoking | 20116 | Smoking | | 421,223 | | *Global P:* | 8·77x10-53 | | *Global P:* | 4·91x10-52 | |
| | 20116 | | Never | | 230,529 (54·7%) | Reference | | | Reference | | |
| | 20116 | | Previous | | 146,351 (34·7%) | -0·022 (-0·029, -0·016) | 1·36x10-11 | -0·96 | -0·022 (-0·029, -0·016) | 1·51x10-11 | -0·97 |
| | 20116 | | Current | | 44,343 (10·5%) | -0·079 (-0·089, -0·069) | 4·71x10-52 | -3·43 | -0·079 (-0·089, -0·068) | 3·15x10-51 | -3·42 |
| | 20161 | Pack years of smoking | | 127,781 | 23·20 (18·01) | -0·035 (-0·041, -0·030) | 4·52x10-35 | -1·52 | -0·027 (-0·032, -0·023) | 1·81x10-14 | -1·18 |
| | 189 | Townsend deprivation index at recruitment | | 422,260 | -1·32 (3·08) | -0·014 (-0·017, -0·011) | 1·46x10-19 | -0·61 | -0·014 (-0·017, -0·011) | 1·37x10-19 | -0·61 |
| | 806 | Standing job | | 242,435 | | *Global P:* | 2·57x10-26 | | *Global P:* | 3·00x10-15 | |
| | 806 | | Never/rarely | | 85,691 (35·3%) | Reference | | | Reference | | |

| | | | | | | | | | | | |
|---|---|---|---|---|---|---|---|---|---|---|---|
| Socioeconomic | 806 | | Sometimes | | 74,453 (30·7%) | -0·020 (-0·030, -0·011) | 2·78x10-05 | -0·87 | -0·014 (-0·023, -0·006) | 0·002 | -0·63 |
| | 806 | | Usually | | 35,773 (14·8%) | -0·028 (-0·040, -0·016) | 5·21x10-06 | -1·22 | -0·022 (-0·033, -0·011) | 1·62x10-04 | -0·96 |
| | 806 | | Always | | 46,518 (19·2%) | -0·062 (-0·073, -0·051) | 4·33x10-28 | -2·70 | -0·045 (-0·054, -0·035) | 2·27x10-15 | -1·94 |
| | 816 | Manual job | | 242,483 | | *Global P:* | 4·93x10-28 | | *Global P:* | 4·39x10-15 | |
| | 816 | | Never/rarely | | 158,031 (65·2%) | Reference | | | Reference | | |
| | 816 | | Sometimes | | 52,058 (21·5%) | -0·038 (-0·048, -0·029) | 5·42x10-15 | -1·65 | -0·033 (-0·042, -0·023) | 8·42x10-09 | -1·42 |
| | 816 | | Usually | | 16,416 (6·8%) | -0·061 (-0·077, -0·045) | 2·29x10-14 | -2·65 | -0·043 (-0·058, -0·028) | 3·51x10-07 | -1·86 |
| | 816 | | Always | | 15,978 (6·6%) | -0·055 (-0·071, -0·040) | 7·59x10-12 | -2·39 | -0·047 (-0·062, -0·032) | 1·05x10-07 | -2·04 |
| | 826 | Shift job | | 242,179 | | *Global P:* | 4·45x10-11 | | *Global P:* | 6·68x10-09 | |
| | 826 | | Never/rarely | | 200,323 (82·7%) | Reference | | | Reference | | |
| | 826 | | Sometimes | | 18,059 (7·5%) | -0·027 (-0·042, -0·012) | 3·57x10-04 | -1·17 | -0·026 (-0·040, -0·012) | 2·98x10-04 | -1·13 |
| | 826 | | Usually | | 5,168 (2·1%) | -0·048 (-0·075, -0·021) | 4·22x10-04 | -2·09 | -0·041 (-0·065, -0·016) | 0·002 | -1·76 |
| | 826 | | Always | | 18,629 (7·7%) | -0·042 (-0·057, -0·028) | 1·40x10-08 | -1·83 | -0·038 (-0·052, -0·024) | 1·55x10-06 | -1·66 |
| | 6138 | Educational qualifications | | 418,349 | | *Global P:* | 2·94x10-154 | | *Global P:* | 1·40x10-152 | |
| | 6138 | | None | | 71,004 (17·0%) | Reference | | | Reference | | |
| | 6138 | | O-levels/CSE | | 70,325 (16·8%) | 0·038 (0·028, 0·048) | 4·64x10-13 | 1·19 | 0·037 (0·027, 0·048) | 1·12x10-12 | 1·63 |
| | 6138 | | A-levels/NVQ/Other | | 138,330 (33·1%) | 0·061 (0·052, 0·070) | 5·96x10-40 | 2·65 | 0·060 (0·051, 0·069) | 3·28x10-39 | 2·62 |
| | 6138 | | Degree | | 138,690 (33·2%) | 0·116 (0·107, 0·125) | 2·40x10-137 | 5·04 | 0·115 (0·106, 0·124) | 4·85x10-136 | 5.00 |
| | 6141 | Marital status | | 419,742 | 307,527 (73·3%) | 0·000 (-0·007, 0·007) | 0·984 | 0.00 | 0·001 (-0·006, 0·007) | 0·817 | 0·03 |
| | 24004 | Air pollution | | 416,537 | 43·84 (14·65) | -0·005 (-0·008, -0·002) | 3·84x10-04 | -0·22 | -0·005 (-0·008, -0·002) | 3·66x10-04 | -0·24 |

Models are adjusted for age, sex, ethnicity and white blood cell count. UKB field, UK Biobank code from which the trait data are derived; N, available sample size for the trait; Values are shown as mean (SD) for continuous traits and as frequencies (%) for categorical variables. Reference indicates the reference category for categorical traits. For categorical variables with more than two categories a Global P has been estimated using a likelihood ratio test and P value given in the P value column. Note that the Bonferonni corrected P value for the number of tests carried out is $4{\cdot}27\text{x}10^{-4}$. Traits that were significant are highlighted in green. Equivalent years of age-related change in LTL is the ratio of the trait beta and the absolute value of age beta (-0·023). Colour coding is to help identify those with effect ≥ 1 year age-related change (in absolute value) in LTL.

Gradient in years of age-related change (in absolute value) in LTL

| 1-1·99 | 2-2·99 | 3-3·99 | 4+ |
|---|---|---|---|

**Supplementary Table 2: Scoring system for the primary and second healthy behaviour indices.**

| Component | UKB field code | Classification | Scoring for | |
|---|---|---|---|---|
| | | | Primary healthy index | Second healthy index |
| Body mass index | 23104 | <18·5 kg/m2 | excluded | 1 |
| | | 18·5-24·9 kg/m2 | 1 | 1 |
| | | 25·0-29·9 kg/m2 | 0 | 1 |
| | | ≥30 kg/$m^2$ | 0 | 0 |
| Smoking | 20116 | Never | 1 | 1 |
| | | Previous | 0 | 1 |
| | | Current | 0 | 0 |
| Diet score | Derived from touchscreen | 0-3 | 0 | 0 |
| | food frequency questionnaire | 4-7 | 1 | 1 |
| Physical activity | 22040 | <735 MET min/week | 0 | - |
| | | ≥735 MET min/week | 1 | - |
| | 22032 | low | - | 0 |
| | | moderate | - | 1 |
| | | high | - | 1 |
| Alcohol intake | Derived from weekly | Female: <5 or >15 g/day | 0 | - |
| | alcohol questionnaire | Male: <5 or >30 g/day | 0 | - |
| | | Female: 5-15 g/day | 1 | - |
| | | Male: 5-30 g/day | 1 | - |

**Supplementary Table 3: Multivariable model results for 17 traits with an association with leucocyte telomere length (LTL) equivalent to ≥ 2 years of age-associated change in LTL.**

| | | | Univariate models | | | Mutlivariable models | | | | | |
|---|---|---|---|---|---|---|---|---|---|---|---|
| | | | Available data (N=84,462) | | | Available data (N=84,462) | | | Imputed data (N=422,797) | | |
| **Group** | **UKB field** | **Trait** | **Beta (95% CI)** | **Pvalue** | **Equivalent years of age-related change in LTL** | **Beta (95% CI)** | **Pvalue** | **Equivalent years of age-related change in LTL** | **Beta (95% CI)** | **Pvalue** | **Equivalent years of age-related change in LTL** |
| Diet | 1329 | Oily fish intake | *Global P:* | 7·34x10-07 | | *Global P:* | 0·001 | | *Global P:* | 4·49x10-08 | |
| | 1329 | Never | Reference | | | Reference | | | Reference | | |
| | 1329 | Less than once a week | 0·033 (0·011, 0·055) | 0·004 | 1·43 | 0·027 (0·005, 0·050) | 0·019 | 1·17 | 0·025 (0·014, 0·035) | 3·47x10-06 | 1·09 |
| | 1329 | Once a week | 0·048 (0·026, 0·070) | 1·87x10-05 | 2·09 | 0·035 (0·012, 0·058) | 0·003 | 1·52 | 0·028 (0·017, 0·038) | 2·11x10-07 | 1·22 |
| | 1329 | 2-4 times a week | 0·071 (0·045, 0·097) | 7·15x10-08 | 3·09 | 0·055 (0·028, 0·082) | 5·53x10-05 | 2·39 | 0·036 (0·024, 0·047) | 3·72x10-09 | 1·57 |
| | 1329 | 5+ times a week | 0·083 (0·004, 0·162) | 0·039 | 3·61 | 0·059 (-0·020, 0·138) | 0·143 | 2·57 | 0·053 (0·022, 0·085) | 9·40x10-04 | 2·30 |
| | 1349 | Processed meat intake | *Global P:* | 1·17x10-04 | | *Global P:* | 0·173 | | *Global P:* | 1·89x10-05 | |
| | 1349 | Never | Reference | | | Reference | | | Reference | | |
| | 1349 | Less than once a week | -0·033 (-0·059, -0·007) | 0·013 | -1·43 | -0·020 (-0·046, 0·007) | 0·152 | -0·87 | -0·016 (-0·027, -0·004) | 0·007 | -0·70 |
| | 1349 | Once a week | -0·052 (-0·078, -0·026) | 1·00x10-04 | -2·26 | -0·031 (-0·058, -0·004) | 0·026 | -1·35 | -0·027 (-0·039, -0·015) | 4·99x10-06 | -1·17 |
| | 1349 | 2-4 times a week | -0·058 (-0·085, -0·032) | 1·78x10-05 | -2·52 | -0·031 (-0·059, -0·004) | 0·027 | -1·35 | -0·027 (-0·039, -0·015) | 8·83x10-06 | -1·17 |
| | 1349 | 5+ times a week | -0·059 (-0·099, -0·019) | 0·004 | -2·57 | -0·031 (-0·072, 0·009) | 0·132 | -1·35 | -0·025 (-0·044, -0·007) | 0·006 | -1·09 |
| | 1408 | Cheese intake | *Global P:* | 7·76x10-10 | | *Global P:* | 1·88x10-05 | | *Global P:* | 2·64x10-08 | |
| | 1408 | Never | Reference | | | Reference | | | Reference | | |
| | 1408 | Less than once a week | -0·011 (-0·058, 0·036) | 0·645 | -0·48 | -0·022 (-0·069, 0·025) | 0·361 | -0·96 | 0·001 (-0·018, 0·021) | 0·888 | 0·04 |
| | 1408 | Once a week | -0·036 (-0·082, 0·010) | 0·128 | -1·57 | -0·045 (-0·091, 0·001) | 0·054 | -1·96 | 0·002 (-0·018, 0·021) | 0·875 | 0·09 |
| | 1408 | 2-4 times a week | -0·010 (-0·055, 0·034) | 0·648 | -0·43 | -0·028 (-0·073, 0·017) | 0·223 | -1·22 | 0·002 (-0·017, 0·021) | 0·839 | 0·09 |
| | 1408 | 5+ times a week | 0·046 (-0·002, 0·093) | 0·060 | 2.00 | 0·015 (-0·033, 0·063) | 0·542 | 0·65 | 0·031 (0·011, 0·051) | 0·002 | 1·35 |
| | 1448 | Bread type | *Global P:* | 8·07x10-11 | | *Global P:* | 0·034 | | *Global P:* | 2·24x10-04 | |
| | 1448 | White | Reference | | | Reference | | | Reference | | |
| | 1448 | Brown | 0·048 (0·025, 0·071) | 2·99x10-05 | 2·09 | 0·030 (0·007, 0·052) | 0·011 | 1·30 | 0·012 (0·002, 0·022) | 0·023 | 0·52 |
| | 1448 | Wholemeal | 0·058 (0·042, 0·074) | 2·05x10-12 | 2·52 | 0·023 (0·005, 0·040) | 0·010 | 1.00 | 0·017 (0·010, 0·025) | 1·08x10-05 | 0·74 |
| | 1448 | Other type | 0·040 (0·003, 0·077) | 0·034 | 1·74 | 0·016 (-0·021, 0·053) | 0·391 | 0·70 | 0·017 (0·001, 0·033) | 0·041 | 0·74 |
| | 1468 | Cereal type | *Global P:* | 3·43x10-13 | | *Global P:* | 0·002 | | *Global P:* | 1·01x10-06 | |
| | 1468 | Other | Reference | | | Reference | | | Reference | | |
| | 1468 | Bran cereal | 0·021 (0·000, 0·042) | 0·055 | 0·90 | -0·001 (-0·023, 0·021) | 0·926 | -0·04 | 0·002 (-0·009, 0·012) | 0·765 | 0·09 |
| | 1468 | Biscuit cereal | 0·005 (-0·016, 0·026) | 0·645 | 0·21 | -0·004 (-0·025, 0·017) | 0·681 | -0·19 | -0·006 (-0·017, 0·004) | 0·245 | -0·26 |
| | 1468 | Oat cereal | 0·034 (0·014, 0·054) | 7·90x10-04 | 1·50 | 0·011 (-0·009, 0·032) | 0·287 | 0·49 | 0·006 (-0·004, 0·017) | 0·249 | 0·26 |
| | 1468 | Muesli | 0·075 (0·055, 0·096) | 8·82x10-13 | 3·27 | 0·034 (0·013, 0·056) | 0·002 | 1·50 | 0·024 (0·013, 0·035) | 1·97x10-05 | 1·04 |
| | 1478 | Added salt | *Global P:* | 3·40x10-04 | | *Global P:* | 0·227 | | *Global P:* | 7·25x10-07 | |
| | 1478 | Never/rarely | Reference | | | Reference | | | Reference | | |
| | 1478 | Sometimes | -0·022 (-0·037, -0·007) | 0·005 | -0·96 | -0·010 (-0·025, 0·005) | 0·187 | -0·43 | -0·013 (-0·020, -0·006) | 2·09x10-04 | -0·57 |
| | 1478 | Usually | -0·016 (-0·039, 0·007) | 0·167 | -0·70 | 0·001 (-0·022, 0·024) | 0·948 | 0·04 | -0·019 (-0·028, -0·009) | 1·24x10-04 | -0·83 |
| | 1478 | Always | -0·068 (-0·106, -0·030) | 4·23x10-04 | -2·96 | -0·034 (-0·072, 0·005) | 0·087 | -1·48 | -0·028 (-0·043, -0·014) | 9·89x10-05 | -1·22 |
| | 2654 | Vegetable spread type | *Global P:* | 2·77x10-04 | | *Global P:* | 0·466 | | *Global P:* | 0·190 | |
| | 2654 | Olive oil based spread | Reference | | | Reference | | | Reference | | |

| | | | | | | | | | | | |
|---|---|---|---|---|---|---|---|---|---|---|---|
| | 2654 | Flora/Benecol | -0·018 (-0·040, 0·004) | 0·107 | -0·78 | -0·010 (-0·033, 0·012) | 0·354 | -0·45 | -0·002 (-0·015, 0·011) | 0·739 | -0·09 |
| | 2654 | Soft (tub) margarine | -0·050 (-0·073, -0·027) | 2·16x10-05 | -2·16 | -0·012 (-0·035, 0·012) | 0·329 | -0·51 | -0·020 (-0·034, -0·005) | 0·007 | -0·87 |
| | 2654 | Hard (block) margarine | 0·003 (-0·197, 0·202) | 0·978 | 0·12 | 0·022 (-0·177, 0·222) | 0·825 | 0·98 | 0·011 (-0·031, 0·052) | 0·607 | 0·48 |
| | 2654 | Polyunsaturated/sunflower | -0·034 (-0·051, -0·017) | 7·40x10-05 | -1·48 | -0·019 (-0·036, -0·002) | 0·027 | -0·83 | -0·008 (-0·019, 0·003) | 0·136 | -0·35 |
| | 2654 | Other low/reduced fat spread | -0·030 (-0·054, -0·005) | 0·017 | -1·30 | -0·008 (-0·032, 0·017) | 0·542 | -0·33 | -0·003 (-0·013, 0·007) | 0·546 | -0·13 |
| | 2654 | Other type of spread | -0·042 (-0·081, -0·003) | 0·036 | -1·83 | -0·025 (-0·064, 0·015) | 0·219 | -1·07 | -0·001 (-0·022, 0·020) | 0·935 | -0·04 |
| Early life and sexual health | 1787 | Maternal smoking around birth | -0·043 (-0·057, -0·028) | 5·51x10-09 | -1·87 | -0·029 (-0·043, -0·014) | 9·14x10-05 | -1·26 | -0·036 (-0·043, -0·029) | 1·55x10-24 | -1·57 |
| General health | 2178 | Overall health status | *Global P:* | 4·35x10-10 | | *Global P:* | 0·003 | | *Global P:* | 2·16x10-07 | |
| | 2178 | Excellent | Reference | | | Reference | | | Reference | | |
| | 2178 | Good | -0·039 (-0·057, -0·021) | 1·31x10-05 | -1·70 | -0·021 (-0·038, -0·003) | 0·024 | -0·91 | -0·012 (-0·020, -0·003) | 0·006 | -0·52 |
| | 2178 | Fair | -0·059 (-0·080, -0·037) | 1·16x10-07 | -2·57 | -0·023 (-0·046, -0·001) | 0·045 | -1·00 | -0·024 (-0·034, -0·014) | 5·70x10-06 | -1·04 |
| | 2178 | Poor | -0·135 (-0·184, -0·086) | 7·31x10-08 | -5·87 | -0·091 (-0·142, -0·041) | 4·11x10-04 | -3·96 | -0·045 (-0·062, -0·027) | 4·18x10-07 | -1·96 |
| | 6149 | Dental problems | | | | | | | | | |
| | 6149_4 | Loose teeth (yes vs no) | -0·041 (-0·078, -0·004) | 0·028 | -1·78 | -0·020 (-0·057, 0·017) | 0·290 | -0·87 | -0·035 (-0·049, -0·020) | 3·25x10-06 | -1·52 |
| | 6149_6 | Dentures (yes vs no) | -0·046 (-0·068, -0·024) | 3·91x10-05 | -2·00 | -0·018 (-0·041, 0·004) | 0·115 | -0·78 | -0·029 (-0·038, -0·021) | 9·20x10-12 | -1·26 |
| Physical activity | 924 | Walking pace | *Global P:* | 6·06x10-07 | | *Global P:* | 0·195 | | *Global P:* | 8·55x10-14 | |
| | 924 | Slow pace | Reference | | | Reference | | | Reference | | |
| | 924 | Average pace | 0·035 (-0·001, 0·071) | 0·055 | 1·52 | 0·015 (-0·022, 0·052) | 0·435 | 0·65 | 0·035 (0·023, 0·048) | 9·10x10-09 | 1·52 |
| | 924 | Brisk pace | 0·067 (0·031, 0·103) | 3·11x10-04 | 2·91 | 0·025 (-0·012, 0·063) | 0·187 | 1·09 | 0·050 (0·037, 0·063) | 3·28x10-14 | 2·17 |
| Smoking | 20116 | Smoking | *Global P:* | 7·19x10-05 | | *Global P:* | 0·045 | | *Global P:* | 8·37x10-17 | |
| | 20116 | Never | Reference | | | Reference | | | Reference | | |
| | 20116 | Previous | -0·012 (-0·026, 0·003) | 0·117 | -0·52 | -0·004 (-0·019, 0·010) | 0·551 | -0·17 | -0·013 (-0·020, -0·006) | 1·08x10-04 | -0·57 |
| | 20116 | Current | -0·056 (-0·081, -0·030) | 1·68x10-05 | -2·43 | -0·033 (-0·058, -0·007) | 0·013 | -1·43 | -0·045 (-0·056, -0·035) | 3·54x10-17 | -1·96 |
| Socioeconomic | 806 | Standing job | *Global P:* | 5·68x10-08 | | *Global P:* | 0·251 | | *Global P:* | 0·159 | |
| | 806 | Never/rarely | Reference | | | Reference | | | Reference | | |
| | 806 | Sometimes | -0·022 (-0·038, -0·006) | 0·008 | -0·96 | -0·011 (-0·027, 0·006) | 0·212 | -0·48 | -0·009 (-0·019, 0·000) | 0·047 | -0·39 |
| | 806 | Usually | -0·046 (-0·066, -0·025) | 1·03x10-05 | -2·00 | -0·023 (-0·045, 0·000) | 0·048 | -1·00 | -0·008 (-0·020, 0·004) | 0·173 | -0·35 |
| | 806 | Always | -0·051 (-0·070, -0·032) | 8·97x10-08 | -2·22 | -0·012 (-0·037, 0·012) | 0·322 | -0·52 | -0·012 (-0·026, 0·001) | 0·075 | -0·52 |
| | 816 | Manual job | *Global P:* | 3·90x10-11 | | *Global P:* | 0·079 | | *Global P:* | 0·789 | |
| | 816 | Never/rarely | Reference | | | Reference | | | Reference | | |
| | 816 | Sometimes | -0·046 (-0·062, -0·030) | 2·51x10-08 | -2·00 | -0·017 (-0·036, 0·002) | 0·082 | -0·74 | -0·005 (-0·016, 0·006) | 0·380 | -0·22 |
| | 816 | Usually | -0·067 (-0·093, -0·041) | 5·23x10-07 | -2·91 | -0·026 (-0·056, 0·005) | 0·096 | -1·13 | -0·006 (-0·025, 0·013) | 0·534 | -0·26 |
| | 816 | Always | -0·039 (-0·066, -0·012) | 0·005 | -1·70 | 0·010 (-0·023, 0·043) | 0·568 | 0·43 | 0·000 (-0·020, 0·020) | 0·999 | 0.00 |
| | 826 | Shift job | *Global P:* | 0·013 | | *Global P:* | 0·539 | | *Global P:* | 0·570 | |
| | 826 | Never/rarely | Reference | | | Reference | | | Reference | | |
| | 826 | Sometimes | -0·034 (-0·060, -0·009) | 0·008 | -1·48 | -0·014 (-0·040, 0·012) | 0·301 | -0·61 | -0·006 (-0·019, 0·008) | 0·413 | -0·26 |
| | 826 | Usually | -0·027 (-0·074, 0·020) | 0·263 | -1·17 | 0·001 (-0·046, 0·048) | 0·961 | 0·04 | -0·013 (-0·037, 0·011) | 0·274 | -0·57 |
| | 826 | Always | -0·025 (-0·049, 0·000) | 0·050 | -1·09 | 0·012 (-0·014, 0·038) | 0·369 | 0·52 | -0·005 (-0·020, 0·010) | 0·502 | -0·22 |
| | 6138 | Educational qualifications | *Global P:* | 3·72x10-30 | | *Global P:* | 1·44x10-09 | | *Global P:* | 1·28x10-38 | |
| | 6138 | None | Reference | | | Reference | | | Reference | | |
| | 6138 | O-levels/CSE | 0·022 (-0·005, 0·049) | 0·108 | 0·96 | 0·005 (-0·022, 0·033) | 0·715 | 0·22 | 0·014 (0·004, 0·025) | 0·007 | 0·61 |

| | | | | | | | | | | | | | |
|---|---|---|---|---|---|---|---|---|---|---|---|---|---|
| 6138 | A-levels/NVQ/Other | | 0·054 (0·029, 0·078) | 1·54x10-05 | 2·35 | | 0·031 (0·006, 0·056) | 0·015 | 1·35 | | 0·030 (0·021, 0·040) | 1·39x10-10 | 1·30 |
| 6138 | Degree | | 0·113 (0·088, 0·137) | 1·70x10-19 | 4·91 | | 0·065 (0·039, 0·092) | 1·63x10-06 | 2·83 | | 0·063 (0·052, 0·073) | 1·03x10-33 | 2·74 |

Results are shown for both the subset of participants (n=84,462) with available data for all 17 traits and for imputed data for all participants. In addition, the univariate results for the subset of participants are also shown to compare with the univariate findings shown in Supplementary Table 1. Models are adjusted for age, sex, ethnicity and white blood cell count. UKB field, UK Biobank code from which the trait data are derived; N, available sample size for the trait; Values are shown as mean (SD) for continuous traits and as frequencies (%) for categorical variables. Reference indicates the reference category for categorical traits. For categorical variables with more than two categories a Global P has been estimated using a likelihood ratio test and P value given in the P value column. The Bonferonni corrected P value for the number of tests carried out is $4·27x10^{-04}$.

Gradient in years of age-related change (in absolute value) in LTL

| | | | |
|---|---|---|---|
| 1-1·99 | 2-2·99 | 3-3·99 | 4+ |

**Supplementary Table 4: Multivariable model results for the association of primary healthy behaviour index with leucocyte telomere length (LTL).**

| | Base model | | | | Adjusted model | | | | Final model | | | |
|---|---|---|---|---|---|---|---|---|---|---|---|---|
| | Available data (N=329,907) | | Imputed data (N=422,797) | | Available data (N=328,229) | | Imputed data (N=422,797) | | Available data (N=306,345) | | Imputed data (N=422,797) | |
| | Beta (95% CI) | Pvalue | Beta (95% CI) | Pvalue | Beta (95% CI) | Pvalue | Beta (95% CI) | Pvalue | Beta (95% CI) | Pvalue | Beta (95% CI) | Pvalue |
| No of healthy behaviours, vs None | | | | | | | | | | | | |
| One | 0·009 (-0·009, 0·026) | 0·326 | 0·013 (-0·002, 0·029) | 0·089 | 0·005 (-0·012, 0·023) | 0·561 | 0·010 (-0·005, 0·025) | 0·198 | -0·002 (-0·020, 0·016) | 0·845 | 0·003 (-0·012, 0·018) | 0·703 |
| Two | 0·030 (0·014, 0·047) | 3·67x10-04 | 0·040 (0·026, 0·054) | 4·72x10-08 | 0·026 (0·009, 0·043) | 0·003 | 0·035 (0·021, 0·049) | 1·18x10-06 | 0·018 (0·000, 0·035) | 0·048 | 0·021 (0·007, 0·035) | 3·24x10-03 |
| Three | 0·055 (0·038, 0·072) | 2·52x10-10 | 0·065 (0·051, 0·079) | 5·33x10-18 | 0·049 (0·032, 0·066) | 2·14x10-08 | 0·060 (0·046, 0·074) | 1·44x10-15 | 0·034 (0·016, 0·052) | 2·39x10-04 | 0·038 (0·024, 0·053) | 3·42x10-07 |
| Four | 0·088 (0·069, 0·107) | 6·61x10-20 | 0·096 (0·080, 0·112) | 6·17x10-29 | 0·082 (0·063, 0·101) | 2·22x10-17 | 0·091 (0·075, 0·108) | 3·36x10-26 | 0·061 (0·041, 0·081) | 2·28x10-09 | 0·062 (0·045, 0·079) | 8·55x10-13 |
| Five | 0·107 (0·077, 0·137) | 5·10x10-12 | 0·114 (0·086, 0·142) | 2·66x10-15 | 0·103 (0·072, 0·133) | 3·89x10-11 | 0·109 (0·081, 0·137) | 3·90x10-14 | 0·072 (0·041, 0·104) | 7·68x10-06 | 0·074 (0·046, 0·103) | 3·39x10-07 |
| Age, per year | -0·023 (-0·024, -0·023) | <1·00x10-300 | -0·023 (-0·024, -0·023) | <1·00x10-300 | -0·023 (-0·024, -0·023) | <1·00x10-300 | -0·023 (-0·024, -0·023) | <1·0x10-300 | -0·023 (-0·023, -0·022) | <1·00x10-300 | -0·023 (-0·023, -0·022) | <1·0x10-300 |
| Males vs Females | -0·175 (-0·181, -0·168) | <1·00x10-300 | -0·174 (-0·180, -0·168) | <1·00x10-300 | -0·171 (-0·177, -0·164) | <1·00x10-300 | -0·170 (-0·176, -0·164) | <1·0x10-300 | -0·234 (-0·261, -0·207) | 7·82x10-65 | -0·239 (-0·262, -0·216) | 2·33x10-84 |
| Ethnicity, vs White | | | | | | | | | | | | |
| Mixed | 0·132 (0·088, 0·176) | 2·99x10-09 | 0·127 (0·089, 0·165) | 7·51x10-11 | 0·133 (0·089, 0·176) | 3·02x10-09 | 0·128 (0·090, 0·166) | 4·55x10-11 | 0·118 (0·072, 0·163) | 3·86x10-07 | 0·126 (0·088, 0·164) | 9·49x10-11 |
| Asian/ Asian British | 0·024 (-0·002, 0·050) | 0·066 | 0·031 (0·010, 0·052) | 0·004 | 0·032 (0·006, 0·058) | 0·016 | 0·039 (0·017, 0·060) | 3·63x10-04 | 0·027 (0·000, 0·054) | 0·054 | 0·036 (0·015, 0·057) | 8·27x10-04 |
| Black/ Black British | 0·389 (0·360, 0·418) | 2·13x10-152 | 0·392 (0·369, 0·416) | 1·57x10-227 | 0·394 (0·365, 0·423) | 5·24x10-154 | 0·395 (0·371, 0·419) | 2·76x10-229 | 0·364 (0·330, 0·398) | 1·35x10-96 | 0·367 (0·340, 0·394) | 1·14x10-154 |
| Chinese | 0·346 (0·286, 0·407) | 4·46x10-29 | 0·365 (0·314, 0·417) | 8·64x10-44 | 0·347 (0·285, 0·408) | 1·38x10-28 | 0·367 (0·315, 0·419) | 3·55x10-44 | 0·322 (0·257, 0·388) | 3·00x10-22 | 0·349 (0·298, 0·401) | 3·23x10-40 |
| Other ethnic group | 0·157 (0·120, 0·194) | 7·44x10-17 | 0·179 (0·148, 0·210) | 2·19x10-30 | 0·159 (0·122, 0·196) | 5·63x10-17 | 0·182 (0·151, 0·213) | 2·24x10-31 | 0·152 (0·113, 0·191) | 2·60x10-14 | 0·174 (0·143, 0·205) | 8·72x10-29 |
| WBC, per one SD higher level | -0·042 (-0·045, -0·039) | 1·41x10-128 | -0·043 (-0·046, -0·040) | 1·98x10-172 | -0·041 (-0·044, -0·037) | 1·71x10-118 | -0·042 (-0·045, -0·039) | 9·86x10-161 | -0·034 (-0·038, -0·031) | 7·84x10-74 | -0·035 (-0·038, -0·032) | 1·76x10-105 |
| Diabetes vs No | - | | - | | -0·051 (-0·067, -0·035) | 2·39x10-10 | -0·047 (-0·061, -0·034) | 1·10x10-11 | -0·028 (-0·045, -0·011) | 0·001 | -0·023 (-0·037, -0·009) | 9·66x10-04 |
| Cancer vs No | - | | - | | 0·006 (-0·007, 0·018) | 0·385 | 0·011 (0·000, 0·022) | 0·056 | 0·008 (-0·005, 0·021) | 0·232 | 0·012 (0·001, 0·024) | 0·030 |
| Hypertension vs No | - | | - | | 0·019 (0·011, 0·027) | 2·26x10-06 | 0·021 (0·014, 0·028) | 1·95x10-09 | 0·031 (0·022, 0·039) | 4·10x10-13 | 0·033 (0·026, 0·040) | 4·79x10-20 |
| Vascular disease vs No | - | | - | | -0·078 (-0·094, -0·063) | 1·61x10-24 | -0·081 (-0·094, -0·068) | 3·12x10-34 | -0·051 (-0·067, -0·035) | 3·30x10-10 | -0·054 (-0·067, -0·041) | 1·93x10-15 |
| Highest qualification, vs None | | | - | | - | | - | | | | | |
| O-level/(G)CSE | - | | - | | - | | - | | 0·024 (0·011, 0·037) | 2·31x10-04 | 0·027 (0·017, 0·038) | 1·79x10-07 |
| A-levels/NVQ/Other | - | | - | | - | | - | | 0·047 (0·036, 0·058) | 2·32x10-16 | 0·048 (0·039, 0·057) | 1·36x10-25 |
| Degree | - | | - | | - | | - | | 0·095 (0·083, 0·106) | 6·21x10-60 | 0·096 (0·086, 0·105) | 5·20x10-91 |
| Suffer from insomnia, vs Never | | | - | | | | - | | | | | |
| Sometimes | - | | - | | - | | - | | -0·003 (-0·011, 0·006) | 0·510 | -0·004 (-0·012, 0·003) | 0·246 |
| Usually | - | | - | | - | | - | | -0·012 (-0·022, -0·002) | 0·017 | -0·011 (-0·019, -0·002) | 0·013 |
| Fed up feelings: Yes versus No | - | | - | | - | | - | | -0·013 (-0·021, -0·006) | 3·96x10-04 | -0·015 (-0·021, -0·008) | 4·90x10-06 |
| LDL, per one SD higher level | - | | - | | - | | - | | 0·024 (0·020, 0·027) | 1·92x10-36 | 0·023 (0·020, 0·026) | 3·22x10-44 |
| CRP, per one SD higher level | - | | - | | - | | - | | -0·015 (-0·019, -0·011) | 2·52x10-14 | -0·017 (-0·020, -0·013) | 4·16x10-23 |
| eGFR, per SD higher level | - | | - | | - | | - | | -0·030 (-0·044, -0·017) | 1·03x10-05 | -0·034 (-0·045, -0·022) | 9·59x10-09 |

Findings are shown for the subset of participants with available data to compute the index (white columns) and for the imputed data in the full cohort (blue shaded columns). The base model includes the primary healthy behaviour index, age, sex, ethnicity and white blood cell (WBC) count, while the adjusted model additionally includes self-reported diseases diagnosed by doctor (diabetes, cancer, hypertension and vascular diseases). The final model, in addition to the parameters included in the adjusted model, is also adjusted for highest educational qualification, suffering from insomnia, fed-up feelings, low-density lipoprotein (LDL), C-reactive protein (CRP) and estimated glomerular filtration rate (CKD-EPI; eGFR). All beta coefficients are for z-standardised LTL with the comparator groups specified in the table.

**Supplementary Table 5: Participant demographics by healthy behaviour groups of the second health behaviour index.**

| n (%) | | Overall 331,658 | None 2,069 (0·62) | One 25,220 (7·60) | Two 96,777 (29·2) | Three 169,664 (51·2) | Four 37,928 (11·4) | Pvalue |
|---|---|---|---|---|---|---|---|---|
| zLTL | | 0·007 (1·00) | -0·06 (0·96) | -0·05 (1·01) | -0·02 (1·00) | 0·03 (1·00) | 0·03 (0·99) | 8·52x10-49 |
| Age, years | | 56·3 (8·06) | 54·3 (7·72) | 55·7 (7·84) | 56·0 (8·01) | 56·4 (8·13) | 57·6 (7·91) | 2·45x10-278 |
| Sex, n (%) | | | | | | | | 6·45x10-136 |
| | Female | 171,770 (51·8) | 895 (43·3) | 11,965 (47·4) | 47,559 (49·1) | 91,369 (53·9) | 19,982 (52·7) | |
| | Male | 159,888 (48·2) | 1,174 (56·7) | 13,255 (52·6) | 49,218 (50·9) | 78,295 (46·2) | 17,946 (47·3) | |
| Ethnicity, n (%) | | | | | | | | 4·95x10-47 |
| | White | 315,856 (95·2) | 1,950 (94·3) | 23,825 (94·5) | 91,526 (94·6) | 162,164 (95·6) | 36,391 (96·0) | |
| | Mixed | 1,939 (0·58) | 18 (0·87) | 181 (0·72) | 665 (0·69) | 917 (0·54) | 158 (0·42) | |
| | Asian | 5,711 (1·72) | 25 (1·21) | 449 (1·78) | 1,794 (1·85) | 2,835 (1·67) | 608 (1·60) | |
| | Black | 4,436 (1·34) | 45 (2·17) | 491 (1·95) | 1,697 (1·75) | 1,839 (1·08) | 364 (0·96) | |
| | Chinese | 1,006 (0·30) | 4 (0·19) | 41 (0·16) | 230 (0·24) | 635 (0·37) | 96 (0·25) | |
| | Other | 2,710 (0·82) | 27 (1·30) | 233 (0·92) | 865 (0·89) | 1,274 (0·75) | 311 (0·82) | |
| White blood cell count, $10^9$ cells/Litre | | 6·83 (1·72) | 8·58 (2·01) | 7·69 (1·93) | 7·16 (1·79) | 6·60 (1·59) | 6·41 (1·54) | <1·00x10-300 |
| Highest education, n (%) | | | | | | | | <1·00x10-300 |
| | None | 46,900 (14·1) | 492 (23·9) | 4,793 (19·1) | 15,593 (16·2) | 21,538 (12·8) | 4,484 (11·9) | |
| | O-levels/CSE/GCSE | 54,143 (16·3) | 414 (20·1) | 4,674 (18·7) | 16,688 (17·4) | 27,032 (16·0) | 5,335 (14·2) | |
| | A-levels/NVQ/Other | 110,429 (33·3) | 688 (33·5) | 8,859 (35·4) | 33,338 (34·7) | 55,793 (33·1) | 11,751 (31·2) | |
| | Degree | 118,081 (35·6) | 463 (22·5) | 6,715 (26·8) | 30,476 (31·7) | 64,312 (38·1) | 16,115 (42·8) | |
| | *Missing* | *2,105 (0·63)* | | | | | | |
| Insomnia, n (%) | | | | | | | | 3·36x10-151 |
| | Never/rarely | 84,046 (25·3) | 418 (20·2) | 5,695 (22·6) | 23,707 (24·5) | 43,982 (25·9) | 10,244 (27·0) | |
| | Sometimes | 157,395 (47·5) | 863 (41·7) | 11,071 (43·9) | 45,327 (46·9) | 82,025 (48·4) | 18,109 (47·8) | |
| | Usually | 90,083 (27·2) | 787 (38·1) | 8,443 (33·5) | 27,699 (28·6) | 43,603 (25·7) | 9,551 (25·2) | |
| | *Missing* | *134 (0·04)* | | | | | | |
| Fed-up feelings, n (%) | | | | | | | | <1·00x10-300 |
| | No | 199,467 (60·1) | 794 (39·1) | 12,056 (48·5) | 53,518 (56·2) | 107,749 (64·5) | 25,350 (67·9) | |
| | Yes | 126,902 (38·3) | 1,239 (60·9) | 12,780 (51·5) | 41,705 (43·8) | 59,203 (35·5) | 11,975 (32·1) | |
| | *Missing* | *5,289 (1·59)* | | | | | | |

| | | | | | | | | |
|---|---|---|---|---|---|---|---|---|
| LDL cholesterol, mmol/L | | | | | | | | 3·95x10-27 |
| | Mean (SD) | 3·55 (0·85) | 3·60 (0·94) | 3·54 (0·90) | 3·57 (0·87) | 3·56 (0·84) | 3·48 (0·83) | |
| | *Missing, n (%)* | *14,333 (4·32)* | | | | | | |
| C-reactive protein, mg/L | | | | | | | | <1·00x10-300 |
| | Mean (SD) | 2·42 (3·59) | 4·94 (4·87) | 4·15 (4·72) | 3·00 (3·92) | 1·98 (3·16) | 1·68 (2·90) | |
| | *Missing, n (%)* | *14,421 (4·35)* | | | | | | |
| eGFR, mg/dL | | | | | | | | 1·74x10-71 |
| | Mean (SD) | 74·5 (75·0) | 61·3 (73·5) | 68·6 (76·1) | 70·7 (75·4) | 77·7 (74·8) | 74·9 (73·6) | |
| | *Missing, n (%)* | *13,912 (4·19)* | | | | | | |
| Diabetes GP, n (%) | | | | | | | | <1·00x10-300 |
| | No | 314,558 (94·8) | 1,794 (87·4) | 22,306 (88·8) | 90,072 (93·3) | 163,759 (96·7) | 36,627 (96·7) | |
| | Yes | 16,441 (4·96) | 259 (12·6) | 2,811 (11·2) | 6,471 (6·70) | 5,637 (3·33) | 1,263 (3·33) | |
| | *Missing* | *659 (0·20)* | | | | | | |
| Cancer GP, n (%) | | | | | | | | 0·009 |
| | No | 306,080 (92·3) | 1,923 (93·3) | 23,244 (92·5) | 89,311 (92·6) | 156,812 (92·6) | 34,790 (91·9) | |
| | Yes | 24,790 (7·47) | 139 (6·74) | 1,882 (7·49) | 7,181 (7·44) | 12,513 (7·39) | 3,075 (8·12) | |
| | *Missing* | *788 (0·24)* | | | | | | |
| Hypertension GP, n (%) | | | | | | | | <1·00x10-300 |
| | No | 243,930 (73·6) | 1,296 (62·8) | 15,271 (60·7) | 66,286 (68·6) | 131,553 (77·6) | 29,524 (78·0) | |
| | Yes | 87,302 (26·3) | 768 (37·2) | 9,907 (39·4) | 30,352 (31·4) | 37,923 (22·4) | 8,352 (22·1) | |
| | *Missing* | *426 (0·13)* | | | | | | |
| Vascular disease GP, n (%) | | | | | | | | 6·06x10-207 |
| | No | 313,050 (94·4) | 1,804 (87·4) | 22,875 (90·9) | 90,356 (93·5) | 162,089 (95·6) | 35,926 (94·9) | |
| | Yes | 18,182 (5·48) | 260 (12·6) | 2,303 (9·15) | 6,282 (6·50) | 7,387 (4·36) | 1,950 (5·15) | |
| | *Missing* | *426 (0·13)* | | | | | | |

Values are shown as mean (SD) for continuous traits and as frequencies (%) for categorical variables. zLTL, Z-standardised leukocyte $\log_e$ telomere length; LDL, low-density lipoproteins; CRP, C-reactive protein; eGFR, estimated glomerular filtration rate (CKD-EPI); diseases are self-reported as diagnosed by doctor. P values are estimated using the Jonckheere-Terpstra test for trend for both continuous and categorical traits.

**Supplementary Table 6: Multivariable model results for the association of second healthy behaviour index with leucocyte telomere length (LTL).**

| | Base model | | | | Adjusted model | | | | Final model | | | |
|---|---|---|---|---|---|---|---|---|---|---|---|---|
| | Available data (N=331,658) | | Imputed data (N=422,797) | | Available data (N=329,962) | | Imputed data (N=422,797) | | Available data (N=307,935) | | Imputed data (N=422,797) | |
| | Beta (95% CI) | Pvalue | Beta (95% CI) | Pvalue | Beta (95% CI) | Pvalue | Beta (95% CI) | Pvalue | Beta (95% CI) | Pvalue | Beta (95% CI) | Pvalue |
| No of healthy behaviours, vs None | | | | | | | | | | | | |
| One | 0·028 (-0·016, 0·071) | 0·213 | 0·030 (-0·004, 0·064) | 0·085 | 0·025 (-0·019, 0·069) | 0·259 | 0·026 (-0·008, 0·060) | 0·136 | 0·019 (-0·027, 0·064) | 0·417 | 0·019 (-0·015, 0·053) | 0·270 |
| Two | 0·045 (0·003, 0·087) | 0·038 | 0·062 (0·028, 0·095) | 3·27x10-04 | 0·040 (-0·003, 0·082) | 0·067 | 0·055 (0·022, 0·089) | 0·001 | 0·027 (-0·017, 0·072) | 0·229 | 0·037 (0·003, 0·070) | 0·031 |
| Three | 0·083 (0·041, 0·125) | 1·21x10-04 | 0·097 (0·064, 0·130) | 9·01x10-09 | 0·077 (0·034, 0·119) | 4·20x10-4 | 0·090 (0·057, 0·123) | 1·05x10-07 | 0·057 (0·012, 0·101) | 0·012 | 0·062 (0·029, 0·095) | 2·68x10-04 |
| Four | 0·117 (0·074, 0·160) | 1·06x10-07 | 0·131 (0·097, 0·165) | 8·97x10-14 | 0·111 (0·068, 0·155) | 5·03x10-07 | 0·124 (0·090, 0·158) | 1·56x10-12 | 0·084 (0·039, 0·129) | 2·91x10-04 | 0·090 (0·056, 0·124) | 2·83x10-07 |
| Age, per year | -0·024 (-0·024, -0·023) | <1·00x10-300 | -0·024 (-0·024, -0·023) | <1·00x10-300 | -0·023 (-0·024, -0·023) | <1·00x10-300 | -0·024 (-0·024, -0·023) | <1·00x10-300 | -0·023 (-0·023, -0·022) | <1·00x10-300 | -0·023 (-0·023, -0·022) | <1·00x10-300 |
| Males vs Females | -0·175 (-0·181, -0·168) | <1·00x10-300 | -0·174 (-0·180, -0·168) | <1·00x10-300 | -0·170 (-0·177, -0·164) | <1·00x10-300 | -0·170 (-0·176, -0·164) | <1·00x10-300 | -0·233 (-0·260, -0·206) | 1·51x10-64 | -0·239 (-0·262, -0·216) | 1·24x10-83 |
| Ethnicity, vs White | | | | | | | | | | | | |
| Mixed | 0·128 (0·085, 0·172) | 6·72x10-09 | 0·126 (0·088, 0·164) | 1·10x10-10 | 0·130 (0·086, 0·173) | 5·69x10-09 | 0·127 (0·089, 0·165) | 6·81x10-11 | 0·116 (0·071, 0·161) | 5·16x10-07 | 0·125 (0·087, 0·163) | 1·28x10-10 |
| Asian/ Asian British | 0·021 (-0·004, 0·047) | 0·103 | 0·028 (0·007, 0·050) | 0·008 | 0·029 (0·003, 0·055) | 0·027 | 0·036 (0·015, 0·057) | 8·45x10-04 | 0·025 (-0·003, 0·052) | 0·078 | 0·034 (0·013, 0·055) | 0·002 |
| Black/ Black British | 0·388 (0·359, 0·417) | 1·04x10-151 | 0·394 (0·370, 0·418) | 4·46x10-229 | 0·392 (0·363, 0·421) | 4·07x10-153 | 0·396 (0·372, 0·420) | 1·83x10-230 | 0·363 (0·329, 0·397) | 2·32x10-96 | 0·367 (0·340, 0·394) | 7·01x10-155 |
| Chinese | 0·344 (0·284, 0·405) | 3·11x10-29 | 0·365 (0·313, 0·416) | 1·10x10-43 | 0·344 (0·283, 0·404) | 1·34x10-28 | 0·366 (0·315, 0·418) | 4·86x10-44 | 0·321 (0·256, 0·385) | 2·18x10-22 | 0·349 (0·298, 0·401) | 3·69x10-40 |
| Other ethnic group | 0·158 (0·122, 0·195) | 2·88x10-17 | 0·176 (0·145, 0·206) | 2·53x10-29 | 0·160 (0·123, 0·197) | 2·06x10-17 | 0·179 (0·148, 0·209) | 2·95x10-30 | 0·155 (0·116, 0·194) | 7·28x10-15 | 0·171 (0·141, 0·202) | 5·76x10-28 |
| WBC, per one SD higher level | -0·041 (-0·045, -0·038) | 4·69x10-122 | -0·042 (-0·045, -0·039) | 2·40x10-158 | -0·040 (-0·043, -0·037) | 6·89x10-113 | -0·041 (-0·044, -0·037) | 2·66x10-148 | -0·034 (-0·037, -0·030) | 1·57x10-71 | -0·034 (-0·037, -0·031) | 1·99x10-99 |
| Diabetes vs No | - | | - | | -0·049 (-0·065, -0·033) | 9·74x10-10 | -0·045 (-0·059, -0·031) | 9·07x10-11 | -0·027 (-0·043, -0·010) | 0·002 | -0·022 (-0·036, -0·008) | 0·002 |
| Cancer vs No | - | | - | | 0·005 (-0·008, 0·017) | 0·467 | 0·011 (-0·001, 0·022) | 0·065 | 0·007 (-0·006, 0·020) | 0·285 | 0·012 (0·001, 0·023) | 0·034 |
| Hypertension vs No | - | | - | | 0·019 (0·011, 0·027) | 2·59x10-06 | 0·021 (0·014, 0·028) | 2·14x10-09 | 0·031 (0·022, 0·039) | 4·24x10-13 | 0·033 (0·026, 0·040) | 6·68x10-20 |
| Vascular disease vs No | - | | - | | -0·081 (-0·096, -0·066) | 6·80x10-26 | -0·082 (-0·095, -0·069) | 5·21x10-35 | -0·053 (-0·069, -0·037) | 6·11x10-11 | -0·055 (-0·068, -0·041) | 7·58x10-16 |
| Highest qualification, vs None | | | - | | - | | - | | | | | |
| O-level/(G)CSE | - | | - | | - | | - | | 0·024 (0·012, 0·037) | 1·76x10-04 | 0·027 (0·017, 0·038) | 2·06x10-07 |
| A-levels/NVQ/Other | - | | - | | - | | - | | 0·048 (0·037, 0·059) | 3·72x10-17 | 0·048 (0·039, 0·057) | 1·53x10-25 |
| Degree | - | | - | | - | | - | | 0·097 (0·085, 0·108) | 1·12x10-62 | 0·096 (0·087, 0·106) | 1·87x10-92 |
| Suffer from insomnia, vs Never | | | - | | | | - | | | | | |
| Sometimes | - | | - | | - | | - | | -0·004 (-0·012, 0·005) | 0·377 | -0·005 (-0·012, 0·003) | 0·223 |
| Usually | - | | - | | - | | - | | -0·013 (-0·023, -0·003) | 0·010 | -0·011 (-0·020, -0·003) | 0·009 |
| Fed up feelings vs No | - | | - | | - | | - | | -0·013 (-0·020, -0·006) | 5·18x10-04 | -0·014 (-0·020, -0·008) | 9·23x10-06 |
| LDL, per one SD higher level | - | | - | | - | | - | | 0·023 (0·020, 0·027) | 5·97x10-36 | 0·023 (0·020, 0·026) | 1·29x10-43 |
| CRP, per one SD higher level | - | | - | | - | | - | | -0·015 (-0·018, -0·011) | 6·28x10-14 | -0·016 (-0·020, -0·013) | 4·69x10-22 |
| eGFR, per one SD higher level | - | | - | | - | | - | | -0·030 (-0·043, -0·016) | 1·50x10-05 | -0·034 (-0·045, -0·022) | 9·88x10-09 |

Findings are shown for the subset of participants with available data to compute the index (white columns) and for the imputed data in the full cohort (blue shaded columns). The base model includes the second healthy behaviour index, age, sex, ethnicity and white blood cell (WBC) count, while the adjusted model additionally includes self-reported diseases diagnosed by doctor (diabetes, cancer, hypertension and vascular diseases). The final model, in addition to the parameters included in the adjusted model, is also adjusted for highest educational qualification, suffering from insomnia, fed-up feelings, low-density lipoprotein (LDL), C-reactive protein (CRP) and estimated glomerular filtration rate (CKD-EPI; eGFR). All beta coefficients are for z-standardised LTL with the comparator groups specified in the table.

**Supplementary Table 7: Bi-directional Mendelian Randomisation analysis of the association between leucocyte telomere length (LTL) and years spent in education (Supplementary Table 7a); smoking initiation (Supplementary Table 7b); and smoking burden (Supplementary Table 7c).**

**a Years spent in education on LTL - 1267 SNPs**

| Method of estimation | Beta (95% CI) | Pvalue | I2 |
|---|---|---|---|
| Inverse-variance weighted | 0·096 (0·080, 0·113) | 3·19x10-30 | 51·4 |
| Egger (for pleiotropy) | - | 0·146 | |
| Weighted median | 0·089 (0·071, 0·108) | 5·36x10-21 | |
| Robust adjusted profile score | 0·101 (0·089, 0·112) | <1·00x10-300 | |

**LTL on years spent in education - 87 SNPs**

| Method of estimation | Beta (95% CI) | Pvalue | I2 |
|---|---|---|---|
| Inverse-variance weighted | 0·005 (-0·020, 0·030) | 0·702 | 63·9 |
| Egger (for pleiotropy) | - | 0·018 | |
| Weighted median | -0·009 (-0·036, 0·018) | 0·497 | |
| Robust adjusted profile score | 0·005 (-0·010, 0·020) | 0·517 | |

**b Initiation of regular smoking on LTL - 374 SNPs**

| Method of estimation | Beta (95% CI) | Pvalue | I2 |
|---|---|---|---|
| Inverse-variance weighted | -0·057 (-0·074, -0·040) | 1·42x10-10 | 52·6 |
| Egger (for pleiotropy) | - | 0·794 | |
| Weighted median | -0·062 (-0·081, -0·043) | 1·03x10-10 | |
| Robust adjusted profile score | -0·060 (-0·072, -0·048) | <1·00x10-300 | |

**LTL on initiation of regular smoking - 89 SNPs**

| Method of estimation | Beta (95% CI) | Pvalue | I2 |
|---|---|---|---|
| Inverse-variance weighted | 0·007 (-0·018, 0·032) | 0·608 | 9·8 |
| Egger (for pleiotropy) | - | 0·326 | |
| Weighted median | -0·005 (-0·042, 0·032) | 0·789 | |
| Robust adjusted profile score | 0·007 (-0·017, 0·031) | 0·589 | |

**c Smoking intensity on LTL - 55 SNPs**

| Method of estimation | Beta (95% CI) | Pvalue | I2 |
|---|---|---|---|
| Inverse-variance weighted | -0·064 (-0·103, -0·026) | 0·001 | 62·2 |
| Egger (for pleiotropy) | - | 0·186 | |
| Weighted median | -0·057 (-0·099, -0·015) | 0·008 | |
| Robust adjusted profile score | -0·066 (-0·090, -0·043) | 3·67x10-8 | |

**LTL on years spent in education - 89 SNPs**

| Method of estimation | Beta (95% CI) | Pvalue | I2 |
|---|---|---|---|
| Inverse-variance weighted | -0·019 (-0·073, 0·036) | 0·502 | 23·9 |
| Egger (for pleiotropy) | - | 0·887 | |
| Weighted median | -0·029 (-0·109, 0·051) | 0·480 | |
| Robust adjusted profile score | -0·019 (-0·067, 0·029) | 0·440 | |

In each table the results are shown for LTL as the outcome and LTL as the predictor with the corresponding number of genetic variants (SNPs) used in each analysis. The results of different MR tests are shown (see Supplementary Methods for explanation of each test).

**Supplementary Table 8: Multivariate model results for the risk of selected diseases per SD longer of leucocyte telomere length (LTL), overall and stratified by number of healthy behaviours.**

| | | Base | | | | Full | | | |
|---|---|---|---|---|---|---|---|---|---|
| Diseases | | Overall | HBI: 0-1 | HBI: 2-3 | HBI: 4-5 | Overall | HBI: 0-1 | HBI: 2-3 | HBI: 4-5 |
| *Not sex specific* | | | | | | | | | |
| **Melanoma** | N | 327,212 | 83,033 | 203,393 | 40,786 | 303,876 | 76,876 | 188,890 | 38,110 |
| | n (%) incident cases | 1,782 (0·54) | 397 (0·48) | 1,148 (0·56) | 237 (0·58) | 1,666 (0·55) | 367 (0·48) | 1,072 (0·57) | 227 (0·60) |
| | HR (95% CI) | 1·12 (1·05, 1·21) | 1·10 (0·95, 1·28) | 1·13 (1·04, 1·24) | 1·10 (0·91, 1·34) | 1·14 (1·06, 1·23) | 1·15 (0·98, 1·34) | 1·13 (1·04, 1·24) | 1·13 (0·93, 1·38) |
| | Interaction P | - | - | 0·693 | 0·701 | - | - | 0·911 | 0·943 |
| **Brain cancer** | N | 329,677 | 83,597 | 204,931 | 41,149 | 306,140 | 77,384 | 190,310 | 38,446 |
| | n (%) incident cases | 623 (0·19) | 176 (0·21) | 377 (0·18) | 70 (0·17) | 586 (0·19) | 163 (0·21) | 356 (0·19) | 67 (0·17) |
| | HR (95% CI) | 1·25 (1·11, 1·41) | 1·29 (1·03, 1·61) | 1·24 (1·07, 1·45) | 1·20 (0·84, 1·71) | 1·26 (1·12, 1·42) | 1·32 (1·05, 1·66) | 1·25 (1·07, 1·46) | 1·18 (0·82, 1·69) |
| | Interaction P | - | - | 0·599 | 0·644 | - | - | 0·469 | 0·528 |
| **Kidney cancer** | N | 329,397 | 83,501 | 204,769 | 41,127 | 305,868 | 77,294 | 190,154 | 38,420 |
| | n (%) incident cases | 1,088 (0·33) | 379 (0·45) | 635 (0·31) | 74 (0·18) | 1,002 (0·33) | 351 (0·45) | 584 (0·31) | 67 (0·17) |
| | HR (95% CI) | 1·20 (1·10, 1·32) | 1·33 (1·15, 1·55) | 1·14 (1·02, 1·29) | 1·21 (0·86, 1·71) | 1·22 (1·11, 1·34) | 1·34 (1·15, 1·57) | 1·15 (1·02, 1·30) | 1·26 (0·88, 1·81) |
| | Interaction P | - | - | 0·084 | 0·298 | - | - | 0·084 | 0·436 |
| **Thyroid cancer** | N | 329,568 | 83,574 | 204,868 | 41,126 | 306,036 | 77,363 | 190,252 | 38,421 |
| | n (%) incident cases | 248 (0·08) | 65 (0·08) | 160 (0·08) | 23 (0·06) | 226 (0·07) | 58 (0·07) | 149 (0·08) | 19 (0·05) |
| | HR (95% CI) | 1·53 (1·27, 1·83) | 1·50 (1·05, 2·13) | 1·51 (1·21, 1·90) | 1·73 (0·95, 3·15) | 1·51 (1·25, 1·82) | 1·51 (1·04, 2·19) | 1·45 (1·14, 1·83) | 2·05 (1·10, 3·84) |
| | Interaction P | - | - | 0·9 | 0·621 | - | - | 0·968 | 0·357 |
| **Sarcoma** | N | 329,673 | 83,595 | 204,938 | 41,140 | 306,126 | 77,383 | 190,310 | 38,433 |
| | n (%) incident cases | 242 (0·07) | 65 (0·08) | 153 (0·07) | 24 (0·06) | 223 (0·07) | 61 (0·08) | 138 (0·07) | 24 (0·06) |
| | HR (95% CI) | 1·24 (1·03, 1·50) | 1·56 (1·10, 2·23) | 1·31 (1·04, 1·66) | 0·47 (0·27, 0·84) | 1·23 (1·01, 1·50) | 1·54 (1·07, 2·22) | 1·31 (1·03, 1·68) | 0·47 (0·26, 0·83) |
| | Interaction P | - | - | 0·445 | 0·002 | - | - | 0·519 | 0·002 |
| **Coronary artery disease** | N | 316,160 | 78,508 | 197,440 | 40,212 | 293,685 | 72,719 | 183,398 | 37,568 |
| | n (%) incident cases | 17,782 (5·62) | 5,825 (7·42) | 10,542 (5·34) | 1,415 (3·52) | 16,401 (5·58) | 5,383 (7·40) | 9,696 (5·29) | 1,322 (3·52) |
| | HR (95% CI) | 0·92 (0·90, 0·94) | 0·94 (0·90, 0·98) | 0·92 (0·90, 0·95) | 0·89 (0·83, 0·97) | 0·94 (0·91, 0·96) | 0·95 (0·92, 0·99) | 0·93 (0·91, 0·96) | 0·90 (0·83, 0·98) |
| | Interaction P | - | - | 0·562 | 0·301 | - | - | 0·414 | 0·228 |
| **Coeliac disease** | N | 328,544 | 83,362 | 204,184 | 40,998 | 305,081 | 77,166 | 189,613 | 38,302 |
| | n (%) incident cases | 891 (0·27) | 217 (0·26) | 543 (0·27) | 131 (0·32) | 817 (0·27) | 200 (0·26) | 496 (0·26) | 121 (0·32) |
| | HR (95% CI) | 0·82 (0·74, 0·90) | 0·86 (0·71, 1·05) | 0·79 (0·69, 0·89) | 0·85 (0·66, 1·11) | 0·80 (0·72, 0·89) | 0·86 (0·70, 1·06) | 0·77 (0·68, 0·88) | 0·84 (0·64, 1·11) |
| | Interaction P | - | - | 0·386 | 0·907 | - | - | 0·326 | 0·985 |
| **Liver cirrhosis** | N | 328,686 | 83,162 | 204,422 | 41,102 | 305,213 | 76,979 | 189,834 | 38,400 |
| | n (%) incident cases | 1,366 (0·42) | 631 (0·76) | 693 (0·34) | 42 (0·10) | 1,247 (0·41) | 578 (0·75) | 631 (0·33) | 38 (0·10) |
| | HR (95% CI) | 0·69 (0·64, 0·75) | 0·73 (0·65, 0·82) | 0·68 (0·61, 0·76) | 0·64 (0·40, 1·01) | 0·72 (0·67, 0·78) | 0·75 (0·66, 0·85) | 0·72 (0·64, 0·80) | 0·70 (0·43, 1·15) |
| | Interaction P | - | - | 0·387 | 0·388 | - | - | 0·594 | 0·46 |
| **Colorectal polyps** | N | 322,398 | 81,101 | 200,730 | 40,567 | 299,429 | 75,083 | 186,441 | 37,905 |
| | n (%) incident cases | 24,066 (7·46) | 7,652 (9·44) | 14,283 (7·12) | 2,131 (5·25) | 22,306 (7·45) | 7,102 (9·46) | 13,214 (7·09) | 1,990 (5·25) |
| | HR (95% CI) | 1·02 (1·00, 1·04) | 1·01 (0·98, 1·05) | 1·03 (1·01, 1·06) | 1·02 (0·96, 1·09) | 1·03 (1·01, 1·05) | 1·03 (0·99, 1·06) | 1·04 (1·01, 1·07) | 1·03 (0·96, 1·10) |
| | Interaction P | - | - | 0·487 | 0·681 | - | - | 0·644 | 0·854 |

| | | | | | | | | | |
|---|---|---|---|---|---|---|---|---|---|
| **Hypotheroidism** | N | 314,247 | 78,940 | 195,560 | 39,747 | 291,822 | 73,098 | 181,597 | 37,127 |
| | n (%) incident cases | 6,153 (1·96) | 1,801 (2·28) | 3,785 (1·94) | 567 (1·43) | 5,673 (1·94) | 1,650 (2·26) | 3,493 (1·92) | 530 (1·43) |
| | HR (95% CI) | 0·90 (0·87, 0·94) | 0·85 (0·79, 0·91) | 0·94 (0·89, 0·98) | 0·87 (0·77, 0·99) | 0·92 (0·88, 0·95) | 0·85 (0·79, 0·92) | 0·95 (0·91, 1·00) | 0·89 (0·78, 1·01) |
| | Interaction P | - | - | 0·025 | 0·684 | - | - | 0·026 | 0·729 |
| **Kidney stones** | N | 325,081 | 82,246 | 202,095 | 40,740 | 301,874 | 76,141 | 187,668 | 38,065 |
| | n (%) incident cases | 3,530 (1·09) | 1,080 (1·31) | 2,156 (1·07) | 294 (0·72) | 3,264 (1·08) | 988 (1·30) | 1,998 (1·06) | 278 (0·73) |
| | HR (95% CI) | 1·07 (1·01, 1·12) | 1·08 (0·99, 1·18) | 1·04 (0·97, 1·10) | 1·32 (1·11, 1·57) | 1·08 (1·03, 1·14) | 1·10 (1·00, 1·21) | 1·04 (0·97, 1·11) | 1·39 (1·17, 1·66) |
| | Interaction P | - | - | 0·324 | 0·071 | - | - | 0·181 | 0·055 |
| **Atopic dermatitis** | N | 320,561 | 81,192 | 199,432 | 39,937 | 297,608 | 75,149 | 185,145 | 37,314 |
| | n (%) incident cases | 582 (0·18) | 146 (0·18) | 361 (0·18) | 75 (0·19) | 554 (0·19) | 141 (0·19) | 344 (0·19) | 69 (0·18) |
| | HR (95% CI) | 0·87 (0·77, 0·98) | 0·91 (0·71, 1·16) | 0·85 (0·73, 0·99) | 0·88 (0·63, 1·25) | 0·86 (0·76, 0·98) | 0·88 (0·69, 1·14) | 0·85 (0·72, 1·00) | 0·89 (0·62, 1·28) |
| | Interaction P | - | - | 0·518 | 0·725 | - | - | 0·791 | 0·825 |
| **Rheumatoeid arthritis** | N | 326,068 | 82,281 | 202,878 | 40,909 | 302,798 | 76,188 | 188,398 | 38,212 |
| | n (%) incident cases | 2,958 (0·91) | 1,041 (1·27) | 1,708 (0·84) | 209 (0·51) | 2,722 (0·90) | 959 (1·26) | 1,566 (0·83) | 197 (0·52) |
| | HR (95% CI) | 0·93 (0·88, 0·98) | 0·97 (0·89, 1·06) | 0·91 (0·85, 0·98) | 0·95 (0·77, 1·16) | 0·96 (0·91, 1·02) | 0·99 (0·89, 1·08) | 0·95 (0·88, 1·02) | 1·01 (0·82, 1·25) |
| | Interaction P | - | - | 0·246 | 0·527 | - | - | 0·375 | 0·933 |
| **Mutliple sclerosis** | N | 328,870 | 83,303 | 204,471 | 41,096 | 305,381 | 77,118 | 189,871 | 38,392 |
| | n (%) incident cases | 284 (0·09) | 97 (0·12) | 161 (0·08) | 26 (0·06) | 270 (0·09) | 90 (0·12) | 154 (0·08) | 26 (0·07) |
| | HR (95% CI) | 1·30 (1·10, 1·55) | 1·27 (0·95, 1·71) | 1·29 (1·03, 1·62) | 1·62 (0·92, 2·85) | 1·33 (1·11, 1·58) | 1·31 (0·96, 1·78) | 1·31 (1·03, 1·65) | 1·61 (0·91, 2·83) |
| | Interaction P | - | - | 1.000 | 0·612 | - | - | 0·917 | 0·678 |
| **Meniere's disease** | N | 328,912 | 83,367 | 204,472 | 41,073 | 305,410 | 77,169 | 189,871 | 38,370 |
| | n (%) incident cases | 357 (0·11) | 97 (0·12) | 219 (0·11) | 41 (0·10) | 332 (0·11) | 92 (0·12) | 200 (0·11) | 40 (0·10) |
| | HR (95% CI) | 1·10 (0·94, 1·28) | 1·04 (0·77, 1·40) | 1·15 (0·94, 1·41) | 0·96 (0·60, 1·52) | 1·11 (0·95, 1·31) | 1·07 (0·78, 1·45) | 1·17 (0·95, 1·44) | 0·96 (0·59, 1·54) |
| | Interaction P | - | - | 0·483 | 0·810 | - | - | 0·591 | 0·712 |
| *Men specific* | | | | | | | | | |
| **Prostate** | N | 156,898 | 41,939 | 96,447 | 18,512 | 145,664 | 38,781 | 89,578 | 17,305 |
| | n (%) incident cases | 6,502 (4·14) | 1,678 (4·00) | 4,048 (4·20) | 776 (4·19) | 6,017 (4·13) | 1,546 (3·99) | 3,742 (4·18) | 729 (4·21) |
| | HR (95% CI) | 1·16 (1·12, 1·21) | 1·20 (1·11, 1·29) | 1·14 (1·09, 1·20) | 1·19 (1·07, 1·32) | 1·15 (1·11, 1·20) | 1·18 (1·10, 1·27) | 1·13 (1·08, 1·19) | 1·20 (1·08, 1·34) |
| | Interaction P | - | - | 0·213 | 0·756 | - | - | 0·411 | 0·796 |
| **Bening prostatic hyperplasia** | N | 151,782 | 40,596 | 93,298 | 17,888 | 140,954 | 37,570 | 86,658 | 16,726 |
| | n (%) incident cases | 10,871 (7·16) | 3,146 (7·75) | 6,583 (7·06) | 1,142 (6·38) | 10,038 (7·12) | 2,897 (7·71) | 6,078 (7·01) | 1,063 (6·36) |
| | HR (95% CI) | 1·11 (1·08, 1·15) | 1·14 (1·08, 1·20) | 1·10 (1·06, 1·14) | 1·16 (1·06, 1·27) | 1·12 (1·09, 1·15) | 1·14 (1·08, 1·20) | 1·10 (1·06, 1·14) | 1·19 (1·09, 1·31) |
| | Interaction P | - | - | 0·197 | 0·851 | - | - | 0·268 | 0·583 |
| *Women specific* | | | | | | | | | |
| **Ovarian cyst** | N | 162,515 | 38,939 | 102,091 | 21,485 | 150,926 | 36,069 | 94,789 | 20,068 |
| | n (%) incident cases | 2,239 (1·38) | 581 (1·49) | 1,394 (1·37) | 264 (1·23) | 2,071 (1·37) | 545 (1·51) | 1,284 (1·35) | 242 (1·21) |
| | HR (95% CI) | 1·10 (1·03, 1·17) | 1·21 (1·07, 1·37) | 1·07 (0·99, 1·16) | 1·04 (0·86, 1·24) | 1·11 (1·04, 1·18) | 1·21 (1·07, 1·37) | 1·09 (1·00, 1·18) | 1·04 (0·86, 1·25) |
| | Interaction P | - | - | 0·074 | 0·108 | - | - | 0·168 | 0·127 |
| **Breast cyst** | N | 165,844 | 39,971 | 104,163 | 21,710 | 154,023 | 37,044 | 96,706 | 20,273 |
| | n (%) incident cases | 493 (0·30) | 132 (0·33) | 295 (0·28) | 66 (0·30) | 452 (0·29) | 122 (0·33) | 269 (0·28) | 61 (0·30) |
| | HR (95% CI) | 1·10 (0·96, 1·26) | 1·21 (0·94, 1·57) | 1·14 (0·96, 1·35) | 0·79 (0·55, 1·14) | 1·12 (0·97, 1·28) | 1·17 (0·89, 1·53) | 1·19 (1·00, 1·42) | 0·76 (0·52, 1·11) |

| | | | | | | | | | |
|---|---|---|---|---|---|---|---|---|---|
| | Interaction P | - | - | 0·628 | 0·053 | - | - | 0·852 | 0·117 |
| **Bening breast lump** | N | 168,144 | 40,520 | 105,589 | 22,035 | 156,160 | 37,542 | 98,046 | 20,572 |
| | n (%) incident cases | 676 (0·40) | 171 (0·42) | 439 (0·42) | 66 (0·30) | 625 (0·40) | 162 (0·43) | 401 (0·41) | 62 (0·30) |
| | HR (95% CI) | 1·14 (1·02, 1·28) | 0·98 (0·78, 1·23) | 1·16 (1·01, 1·33) | 1·56 (1·09, 2·24) | 1·16 (1·03, 1·31) | 1·04 (0·82, 1·31) | 1·15 (1·00, 1·33) | 1·64 (1·14, 2·37) |
| | Interaction P | - | - | 0·928 | 0·013 | - | - | 0·377 | 0·024 |
| **Uterine polyps** | N | 163,956 | 39,482 | 102,867 | 21,607 | 152,278 | 36,574 | 95,528 | 20,176 |
| | n (%) incident cases | 3,723 (2·27) | 980 (2·48) | 2,370 (2·30) | 373 (1·73) | 3,443 (2·26) | 905 (2·47) | 2,189 (2·29) | 349 (1·73) |
| | HR (95% CI) | 1·16 (1·10, 1·22) | 1·24 (1·13, 1·36) | 1·12 (1·06, 1·19) | 1·22 (1·05, 1·42) | 1·17 (1·11, 1·23) | 1·26 (1·14, 1·39) | 1·13 (1·06, 1·20) | 1·20 (1·03, 1·41) |
| | Interaction P | - | - | 0·143 | 0·937 | - | - | 0·121 | 0·873 |
| **Uterine fibroid** | N | 158,962 | 38,160 | 99,791 | 21,011 | 147,673 | 35,360 | 92,693 | 19,620 |
| | n (%) incident cases | 3,130 (1·97) | 779 (2·04) | 1,997 (2·00) | 354 (1·68) | 2,917 (1·98) | 738 (2·09) | 1,855 (2·00) | 324 (1·65) |
| | HR (95% CI) | 1·28 (1·22, 1·35) | 1·31 (1·18, 1·46) | 1·29 (1·21, 1·37) | 1·20 (1·02, 1·40) | 1·30 (1·23, 1·37) | 1·31 (1·18, 1·46) | 1·31 (1·23, 1·40) | 1·20 (1·02, 1·42) |
| | Interaction P | - | - | 0·640 | 0·308 | - | - | 0·716 | 0·234 |

The base model is adjusted for age, sex, ethnicity and white blood cell count, whereas the full model is additionally adjusted for self-reported diseases diagnosed by doctor (diabetes, cancer, hypertension, vascular disease), educational level, insomnia, fed-up feelings, and low-density cholesterol, C-reactive protein, estimated glomerular filtration rate (CKD-EPI). N, available sample size; n, number of cases. The P value of the interaction term between the telomere length and the number of healthy behaviours is given in the "Interaction P" row. The selected diseases were those previously identified (Codd V, et al. *Nat Gen* . 2021) to be directionally concordant between mendelian randomisation and observational data analyses at Bonferroni (4·10x10-04) or nominal (5·00x10-02) significance level. In this analysis we examined 22 diseases during two modelling procedures, therefore the Bonferroni significance level was set at 1·14x10-03.
